# A reinforcement learning model to inform optimal decision paths for HIV elimination^1^

**DOI:** 10.1101/2021.07.11.21260328

**Authors:** Seyedeh N. Khatami, Chaitra Gopalappa

**Author notes:** Co-author. Financial support for this study was provided partially by a grant from National Institute of Allergy and Infectious Diseases of the National Institutes of Health under Award Number R01AI127236. The funding agreement ensured the authors’ independence in designing the study, interpreting the data, writing, and publishing the report.

## Abstract

The ‘Ending the HIV Epidemic (EHE)’ national plan aims to reduce annual HIV incidence in the United States from 38,000 in 2015 to 9,300 by 2025 and 3,300 by 2030. Diagnosis and treatment are two most effective interventions, and thus, identifying corresponding optimal combinations of testing and retention-in-care rates would help inform implementation of relevant programs. Considering the dynamic and stochastic complexity of the disease and the time dynamics of decision-making, solving for optimal combinations using commonly used methods of parametric optimization or exhaustive evaluation of pre-selected options are infeasible. Reinforcement learning (RL), an artificial intelligence method, is ideal; however, training RL algorithms and ensuring convergence to optimality are computationally challenging for large-scale stochastic problems. We evaluate its feasibility in the context of the EHE goal.

We trained an RL algorithm to identify a ‘sequence’ of combinations of HIV-testing and retention-in-care rates at 5-year intervals over 2015-2070, which optimally leads towards HIV elimination. We defined optimality as a sequence that maximizes quality-adjusted-life-years lived and minimizes HIV-testing and care-and-treatment costs. We show that solving for testing and retention-in-care rates through appropriate reformulation using proxy decision-metrics overcomes the computational challenges of RL. We used a stochastic agent-based simulation to train the RL algorithm. As there is variability in support-programs needed to address barriers to care-access, we evaluated the sensitivity of optimal decisions to three cost-functions.

The model suggests to scale-up retention-in-care programs to achieve and maintain high annual retention-rates while initiating with a high testing-frequency but relaxing it over a 10-year period as incidence decreases. Results were mainly robust to the uncertainty in costs. However, testing and retention-in-care alone did not achieve the 2030 EHE targets, suggesting the need for additional interventions. The results from the model demonstrated convergence. RL is suitable for evaluating phased public health decisions for infectious disease control.

## 1. Introduction

The human immunodeficiency virus (HIV) continues to persist as a major public health issue in the United States (US), with about 1.2 million people living with HIV (PWH) as of 2015 and about 38,000 becoming newly infected each year [1]. The 2019 ‘Ending the HIV Epidemic (EHE)’ US national strategic plan aims to reduce new infections by about 75% (to 9300 cases) by 2025 and by about 90% (to 3000 cases) by 2030 [2, 3], by scaling-up four strategies, diagnose, treat, prevent, and respond [2]. Diagnoses followed by antiretroviral therapy (ART) treatment are key interventions as, in addition to being therapeutic, they can reduce HIV transmissions by up to 100% [4]. Since the implementation of the first national strategic plan in 2010, national guidelines have recommended at least annual testing for high-risk populations [5] and treatment initiation immediately upon diagnosis. However, estimates from the National HIV Surveillance System (NHSS) indicate that actual testing is less frequent than recommended, e.g., 3 to 5 years among those diagnosed with HIV in 2015 [6, 7]. Further, though an estimated 70% to 80% of persons diagnosed with HIV were linked to care-and-treatment upon diagnosis, only 48% were on ART treatment in 2015, indicating high rates of care drop-out [8].

In this study, we develop a model to identify an optimal sequence of combinations of testing and retention-in-care rates at every 5-year interval from 2015 to 2070, to reduce HIV incidence. Identifying optimal testing rates, which is the inverse of how often to test, helps inform testing guidelines and implement social support and outreach programs to enable uptake [9]. Identifying optimal retention-in-care rates, which is the proportion of persons in care at the end of the year from among those in care at the beginning of that year, helps inform social service and support programs necessary to reduce the current high rates of care drop-out [10]. Identifying risk-group specific testing and retention-in-care rate combinations help direct resources to relevant support programs. We use non-linear cost functions to assign fixed and variable costs of testing and retention-in-care. To model the need for additional outreach and social support services to address barriers to testing and sustained care-and-treatment [11, 12, 13], we assume that the variable unit costs increase non-linearly with testing and retention-in-care rates [14, 15]. As the type of service program-needs and its effectiveness vary by population [16], we utilize varying cost functions to generate the uncertainty range in optimal decisions.

Previous studies that have evaluated combinations of testing and retention-in-care rates have identified the most cost-effective combinations through either use of comparative analysis to evaluate a few pre-selected scenarios in stochastic simulation models [17-19], use of static parametric optimization techniques to evaluate non-dynamic decisions suitable for short-term decision-making [20, 21], or use of dynamic optimal control techniques that evaluate dynamic decisions but using deterministic differential equation-based models [20-22]. Our approach evaluates dynamic decision sequences in a stochastic and dynamic agent-based simulation environment through formulating the problem as a Markov decision process (MDP) and solving using reinforcement learning (RL), an area of artificial intelligence [23-25]. This methodology enables both simulating the dynamic changes in the epidemic over time while also evaluating the corresponding dynamic changes in decisions over time in a stochastic environment to identify the most optimal sequence choices to reduce new infections. We use a previously validated Progression and Transmission of HIV (PATH 2.0) model [26], a dynamic stochastic agent-based model, for the simulation of the epidemic and evaluation of the decisions. Previous RL models in HIV have focused on patient-level clinical decisions such as optimal treatment protocols [27, 28].

Recent literature has seen an emergence in the use of RL for public health decision-making related to the COVID-19 pandemic, but they predominantly use deterministic equation-based model environments [29-34]. While deterministic models are suitable for diseases that spread easily, agent-based models are more suitable for capturing the individual-level interactions for slow-spreading diseases such as HIV [35]. However, a combination of agent-based models and RL create computational challenges. The number of iterations of RL training needed to ensure convergence increases exponentially with the size of the possible choices (225^11^ possible sequences in this application), and agent-based models are computationally expensive (e.g., each iteration of PATH 2.0 takes about 30 minutes). To overcome this challenge, we reformulate the decision variables to use proportions unaware and on ART as proxies, proving it mathematical viability, which reduces the number of choices to 36^11^. To the best of our knowledge, this is the first model to evaluate the EHE goal of HIV elimination as a sequential decision-making problem in a stochastic dynamic environment and is naturally suited for informing the sequential goals of the US national plan.

## 2. Methodology

A MDP is a stochastic formulation of a decision-making problem, and RL is a machine learning methodology that uses 1) a simulation model to evaluate a *policy* (sequence of decisions) and 2) a control optimization algorithm to control the selection of policies to evaluate [36]. We used the Progression and Transmission of HIV/AIDS (PATH 2.0), a stochastic dynamic agent-based simulation model [26], and Q-learning RL algorithm [36, 37]. We describe the MDP formulation in section 2.1, and the RL algorithm in section 2.22.

### 2.1 Mathematical formulation of the decision-making problem as a MDP

Let epidemic *state* at time *t* be a multivariate parameter *X*_*t*_ = [*p*_*i*_, *μ*_*u,i*_, *μ*_*a,i*_, *μ*_*ART,i*_; ∀*i*]_*t*_, where

- *p*_*i*_ is the HIV prevalence calculated as the number of people living with HIV (PWH) in risk group *i* divided by the total number of people in the population; we modeled two risk groups, heterosexuals (HETs) and men who have sex with men (MSM), thus *i* ∈ { HETs, MSM}, and
- *μ*_*u,i*_, *μ*_*a,i*_, *μ*_*ART,i*_ are the proportions of PWH in risk group *i* that are unaware of their infection, aware of their infection but not on ART, and aware and on ART, respectively, and *μ*_*u,i*_ + *μ*_*a,i*_ + *μ*_*ART,i*_ = 1; ∀*i*, thus representing all stages along the care-continuum.

Note that, for HIV prevalence (*p*_*i*_), we use the total population size as the denominator instead of the commonly used public health definition that uses the population size in that specific risk group *i* as the denominator. This modification makes the state space mutually exclusive and collectively exhaustive, a necessary property for a MDP.

Let the intervention decision at time *t* be a multivariate parameter *D*_*t*_ = [*δ*_*i*_, (1 − *ρ*_*i*_); ∀*i*]_*t*_, where

*δ*_*i*_ is the diagnostic rate and

(1 − *ρ*_*i*_) is the retention-in-care rate in risk group *i* where *i* ∈ { HETs, MSM}.

Then, {*X*_*t*_, *D*_*t*_: *t* = 0: T = 2015, 2020, 2025, …, 2070} is a MDP defined with a 4-tuple {Ω, A, *P*_*a*_, *R*_*a*_}, where,

- Ω is the state space, defined as the set of all possible states of the epidemic, i.e., Ω = {[*p*_*i*_, *μ*_*u,i*_, *μ*_*a,i*_, *μ*_*ART,i*_; ∀*i* ∈ { HETs, MSM}]} and *X*_*t*_ ∈ Ω,
- *A* is the action space, defined as the set of all possible decisions (referred to as actions in MDP terminology), i.e., *A* = {[*δ*_*i*_, (1 − *ρ*_*i*_), ∀*i* ∈ {HETs, MSM}]}, and *D*_*t*_ ∈ A,
- *P*_*a*_ is the one-step transition probability matrix under action *a*, with element *P*_*a*_(*x, x*′) being the probability that the epidemic transitions from state *X*_*t*_ = *x* to *X*_*t+1*_ = *x*′ when action *a* is taken, and
- *R*_*a*_ is the immediate reward matrix under action *a*, with element *R*_*a*_(*x, x*′) being the immediate reward (total benefits minus total costs) of taking action *a* when the epidemic is in state *x* and, as a result, it transitions to state *x*′; we model costs as intervention costs and benefits as the total quality-adjusted life-years (QALYs) lived in the population.

Note that the epidemic at any time *t* can be represented by one and only one *state*, and the probability of transitioning to an epidemic state *x*′ at time *t* + 1 is only dependent on the epidemic state *x* at time *t*, i.e., *Pr*[*X*_*t+1*_ *X*_*t*_, *X*_*t−1*_, *X*_*(t−2)*_, …} = *Pr*{*X*_*t+1*_|*X*_*t*_}, thus satisfying the necessary Markov property for the MDP. Also, note that we use *t* = [0: T] = [2015, 2020, 2025, …, 2070] to denote that we evaluate decisions at every five-year interval (consistent with the decisions made in the EHE national strategic plan). The initial year is *t* = 0 = *year* 2015, and thus, the first decision-making interval is for the period 2016 to 2020. We chose 2015 as the start year because, at the time of model development, the latest surveillance data available for HIV was 2016.

The objective is to identify the optimal decision ***d*** ∈ [*d*_*1*_, . ., *d*_*T*_] (referred to as an optimal *policy* in MDP terminology) that maximizes the expected reward, i.e.,

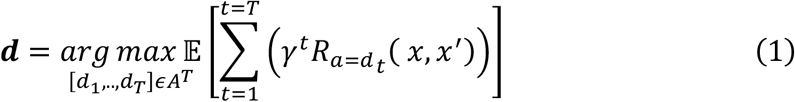

where, *γ* is the discounting factor, and 𝔼[.] is the expected value. Thus, ***d*** is the sequence of optimal actions at 5-years intervals over the period 2016 to 2070. Conceptually, (1) suggests that the decision *d*_*t*_ at every decision-making epoch *t* is evaluated not just based on its costs and impacts during the current epoch 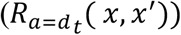, but is also based on the costs and impacts of decisions that would have to be made in all future decision epochs ([*d*_*t+1*_, *d*_*t+2*_, … *d*_*T*_]) to eliminate HIV, while also optimizing those future decisions. Intuitively, a policy that leads to zero new infections will be optimal if it has the lowest future costs and the highest benefits (QALYs), though it may have higher immediate costs. Under this objective function, it is necessary to not discount future costs and benefits as discounting would diminish the weight given to infections averted and costs prevented in the future, and thus will not identify strategies that lead to HIV elimination. Therefore, we set *γ* = 1. The problem of solving for the optimal policy ***d*** can be visualized as a decision-tree (Appendix section 1 Figure A1), with the *epidemic state* in 2015 being the start node, *actions* (*a*) being the decision nodes, *epidemic state* the decision transitions it to in next 5-year interval being the chance nodes (with the probability of transition defined by *P*_*a*_ and value of the outcome defined by *R*_*a*_), the possible *epidemic states* in 2070 being the end nodes, and a *policy* being a path or a sequence of decisions in the decision-tree. Analytically estimating the outcome of each decision path is complex, the dynamics of the system makes estimating *P*_*a*_ and *R*_*a*_ complex, and the large dimensions of the state space and action space, as will be seen in below formulation of the 4-tulpe {Ω, A, *P*_*a*_, *R*_*a*_}, makes it impractical. Therefore, we solve for the optimal policy ***d*** using RL (see section 2.2).

We next discuss the formulation of each element of the 4-tulpe {Ω, A, *P*_*a*_, *R*_*a*_}.

#### State space

We formulate the state space Ω = [[*p*_*i*_, *μ*_*u,i*_, *μ*_*a,i*_, *μ*_*ART,i*_; ∀*i* ∈ { HETs, MSM}]} as a finite state space by discretizing its elements as follows:

Ω = {[*p*_*HET*_, *μ*_*u,HET*_, *μ*_*a,HET*_, *μ*_*ART,HET*_, *p*_*MSM*_, *μ*_*u,MSM*_, *μ*_*a,MSM*_, *μ*_*ART,MSM*_], Δ}, where,

*p*_*HET*_ ∈ {[0,0.005), [0.0005,0.0015), [0.015,0.0025), [0.0025,0.0035), [0.0035,0.0045)}, i.e., all states with HET prevalence in range 0 *to* 0.005 will be assigned *state* = [0,0.005)

*p*_*MSM*_ ∈ {[0.0005,0.0015), [0.015,0.0025), [0.0025,0.0035), …, [0.0085,0.0095), [0.0095,0.0015)}, i.e., all states with MSM prevalence in range 0 *to* 0.005 will be assigned *state* = [0,0.005)

*μ*_*u,HET*_ ∈ {[8.75%, *11*.25%), [*11*.25%, 13.75%), [13.75%, 16.25%)}

*μ*_*u,MSM*_ ∈ {[8.75%, *11*.25%), [*11*.25%, 13.75%), [13.75%, 16.25%), [16.25%, 18.75%)}

*μ*_*ART,i*_ ∈ {[45%, 55%), [55%, 65%), [65%, 75%), [75%, 85%), [85%, 95%)},

*μ*_*a,i*_ = 1 − *μ*_*u,i*_ − *μ*_*ART,i*_, and

Δ is a *state* that includes all other values (i.e., *p* _*HETs*_ ≥ 0.0045, *p* _*MSM*_ ≥ 0.015, *μ*_*u,i*_ < 8.75%, *μ*_*u,HETs*_ ≥ 16.25%, *μ*_*u,MSM*_ ≥ 18.75%, *μ*_*ART,i*_ < 45% *or μ*_*ART,i*_ ≥ 95%).

State Δ is representative of all epidemic states we want to avoid and is allocated a very large cost, such that any action that would take the epidemic to that state would have a high negative reward and thus be marked as a bad decision. The upper bounds on *p*_*i*_, *μ*_*u,i*_ and lower bound on *μ*_*ART,i*_ are set to values in 2015 to indirectly constrain the decisions to lead to a better epidemic state. Considering all the possible combinations of the discretized values, noted above for *p*_*HET*_, *p*_*MSM*_, *μ*_*u,HET*_, *μ*_*u,MSM*_, *μ*_*ART,HET*_, *μ*_*ART,MSM*_, there are a total of 16,500 (= 5 × *11* × 3 × 4 × 5 × 5) *states*. Hence, the final size of the state space is |Ω| = 16,500 + 1.

#### Action space

Instead of directly formulating an action as a combination of diagnostic rate (δ_*i*_) and retention-in-care rate (1 − *ρ*_*i*_), which is the decision of interest here, we formulate it using changes in proportions unaware and on ART as a proxy, i.e., instead of using action space as A = {[*δ*_*HET*_, *δ*_*MSM*_, (1 − *ρ*_*HET*_), (1 − *ρ*_*MSM*_)]} we use a proxy as

*A* = {[*a*_*unaware,HET*_, *a*_*unaware,MSM*_, *a*_*ART,HET*_, *a*_*ART,MSM*_]}, where,

*a*_*unaware,i*_ is the percentage decrement in *μ*_*u,i*_ (the proportion unaware in risk group *i*), and *a*_*ART,i*_ is the percentage increment in *μ*_*ART,i*_ (the proportion on ART in risk group *i*).

We formulated the action space as above because of its attractive mathematical properties that help efficiently constrain the number of action choices and thus improve the chance of convergence of the RL algorithm. We discuss these mathematical properties through four Remarks as follows.

#### Remark 1

Given the system state *x* at time *t* − 1, (*X*_*t−1*_ = *x*), corresponding to every action *a*_*unaware,i*_, there is a unique diagnostic rate (δ_*i*_) and corresponding to every action *a*_*ART,i*_, there is a unique retention-in-care rate (1 − *ρ*_*i*_).

#### Remark 2

From a public health perspective, all actions that transition the epidemic state to a higher proportion unaware (*μ*_*u,i*_) or to a lower proportion on ART (*μ*_*ART,i*_), compared to its current state, are undesirable and should not be selected.

#### Remark 3

Setting the action space to use *a*_*unaware,i*_ and *a*_*ART,i*_ instead of diagnostic rate and retention-in-care rate, respectively, helps efficiently control the number of possible actions (interventions) and thus is more computationally efficient.

#### Remark 4

Setting the action space to use *a*_*unaware,i*_ and *a*_*ART,i*_ instead of *changes* in diagnostic rate and retention-in-care rate, respectively, over two consecutive decision-making time-steps, also helps efficiently control the number of possible actions and thus is more computationally efficient.

We support Remarks 1 to 4 through proofs in the Appendix section 3. Briefly, for any given epidemic state, corresponding to every combination of *a*_*unaware,i*_ and *a*_*ART,i*_, there is a unique combination of diagnostic and retention-in-care rates, which essentially implies that our formulation of *action* as A = [[*a*_*unaware,i*_, *a*_*ART,i*_; ∀*i*]} would yield the same results as the more direct metrics of A = {[*δ*_*i*_, (1 − *ρ*_*i*_); ∀*i*]} (Remark 1). In fact, for evaluating the proxy action in the simulation, we first estimate the diagnostic and retention-in-care rates (see Appendix section 2) and use that as input to the simulation. We use this estimation method, which generates functional expressions between *δ*_*i*_ and *a*_*unaware,i*_ and between (1 − *ρ*_*i*_) and *a*_*ART,i*_ through the representation of the system as a differential equations model (see Appendix section 2), followed by showing that the functional expressions are bijection functions (see Appendix section 3) to prove Remark 1. Note that the proxy action space elements (*a*_*unaware,i*_ and *a*_*ART,i*_) directly modify part of the state space elements (*μ*_*u,i*_ and *μ*_*ART,i*_, respectively), which gives the flexibility to, at any decision time-step, not choose actions that would take the epidemic to a state worse than the current (Remark 2), and thus, also avoid transitioning into the undesirable state Δ. If on the other hand, we choose *δ*_*i*_ and (1 − *ρ*_*i*_) as elements of the action space, we prove that we have to run the simulation model (∼30 mins per run) to evaluate what state that action would transition the system into (could be better than current, worse than current, or Δ) thus requiring many more evaluations (Remark 3). Remark 4 has a similar purpose as Remark 3 except that it evaluates the use of changes in the diagnostic rate and retention-in-care rate over two consecutive decision-making time-steps, as the proxies *a*_*unaware,i*_ and *a*_*ART,i*_ are also decrements or increments (of *μ*_*u,i*_ or *μ*_*ART,i*_, respectively) over two consecutive decision-making time-steps.

We assumed two possible choices for decrements in *μ*_*u,i*_, decrease by 0% or 2.5%, and three possible choices for increments in *μ*_*ART,i*_, increase by 0%, 10%, or 20%, each relative to their values at the time of decision-making. That is, we formulated the possible action choices as

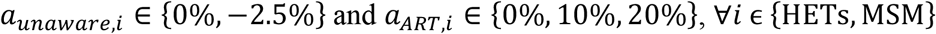

resulting in 36 possible *actions* (2 × 3 for HETs × 2 × 3 for MSM) to choose from at every 5-year decision interval between 2015-2070, and thus, 36^11^ possible decision sequences. If, on the other hand, we had directly formulated an action as changes in testing and retention-in-care rates, we would in the least have 225 action choices, and thus 225^11^ possible sequences (see Appendix section 3). The size of the action space can exponentially increase the number of RL iterations for convergence (see convergence discussion in next section) and thus becomes infeasible to model or guarantee convergence.

For public health decision-making and implementation, testing rates and retention-in-care rates are more meaningful. Therefore, in Results, we present both metrics, the changes in proportions unaware and on ART, and the direct metrics of testing rate, estimated from the simulation as the inverse of the time from infection to diagnosis (a proxy for how often to test), and retention-in-care rate, estimated from the simulation as the proportion retained in care for the entire year among those in care at the beginning of that year.

#### Transition probabilities and immediate rewards

Generating the full one-step transition probability matrices *P*_*a*_ and reward matrices *R*_*a*_ is infeasible considering the size of the state space and action space. Therefore, we used PATH 2.0 [26], discussed later under Simulator, to simulate *actions* and stochastic *transitions*, track corresponding *states* it transitions to, and estimate *immediate rewards*. We estimated the *immediate rewards* as benefits minus costs. We model benefits as the total population quality-adjusted-life-years (QALYs) lived converted to a monetary value by multiplying with the US gross domestic product (GDP) per capita of $54,000 to denote the economic value added for every QALY lived [9, 38, 39]. Costs include total population costs for HIV testing, care, and treatment. Specifically, we estimated *R*_*a*_(*x, x*′) = (*c*_*l*_*L*_*t*_ − *C*_*t*_), where,

*c*_*l*_ = cost per QALY lived, a health utility measure to control for the willingness to pay for every QALY lived; here we assumed it is equal to $54,000, the GDP per capita in the US in year 2015,

*L*_*t*_ = sum of QALYs of all people in the population at decision-making epoch *t*,

*C*_*t*_ = sum of HIV-related costs and intervention costs at decision-epoch *t*.

We used the PATH 2.0 simulation model for the estimation of *C*_*t*_ and *L*_*t*_. We present the estimation of the unit-costs corresponding to the interventions in Appendix section 4.

#### Constraints

We set the following constraints, which can be interpreted as cost or feasibility constraints:

a. The maximum possible decrement in proportion unaware and the maximum possible increment in proportion on ART, achievable in a 5-year interval, were set at 2.5% and 20%, respectively, as evident from the choice of *actions*. For reference as to the feasibility of achieving these maximum scale-ups, between 2010 and 2015, there was about a 2.3% decrease in proportion unaware (17.3% in 2010 to 15% in 2014) and a 13.8% increase in proportion on ART (46% in 2010 to 59.8% in 2015) [40, 41].
b. The maximum proportion aware and maximum proportion on ART were set at about 95%, which are the targets typically aimed in national and global strategic plans [2, 42].

### 2.2 Reinforcement learning algorithm to identify optimal policy

Reinforcement learning (RL) and dynamic programming (DP) are commonly used algorithms to solve MDP problems. Applying DP guarantees convergence to the optimal solution; however, it requires estimating, under each action, a probability matrix of transitions between all states. In the case of large-scale problems such as our current application to HIV, estimating the transition probability matrices for all states and actions is computationally infeasible. This curse of dimensionality makes DP suitable for only small-scale problems [36]. Therefore, we use Q-learning, a machine learning control optimization algorithm that uses an iterative feedback and control process to identify the optimal policy. Thus, it does not require a priori knowledge of transition probability matrices and is known to converge to near-optimal solutions [36].

#### Q-learning algorithm

In this study, we use PATH 2.0 (‘simulator’) to simulate a specific *action* and train the Q-learning algorithm. The simulator returns as ‘feedback’ to the Q-learning algorithm (‘optimizer’), the *immediate reward* of the action simulated, and the epidemic *state* it transitions to at the end of the 5-year period. The optimizer tracks the *total reward* for the period 2015 to 2070 by summing the *immediate reward* of each 5-year *action* while also tracking the epidemic states visited. It then controls what action should be next taken by observing the total reward of previous actions and sending that decision back to the simulator [36, 43, 44] . By repeating this iterative process a large number of times (training the Q-learning algorithm), the optimizer learns to pick a better action each time to eventually find the optimal decision.

The optimizer tracks total reward as Q-values,

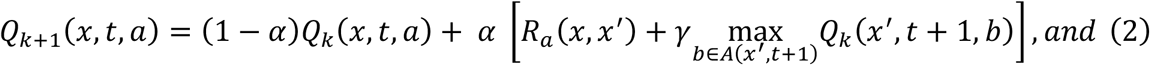

at every decision epoch *t*, given system state as *x*, selects the action (*a*(*x, t*)) to simulate using a decaying epsilon greedy method,

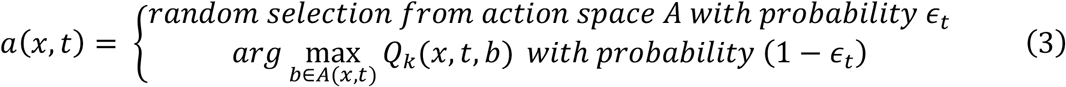

where *k* is the iteration of the Q-learning algorithm, *α* is a learning rate, *γ* is the discounting factor, which was set to 1 here, and 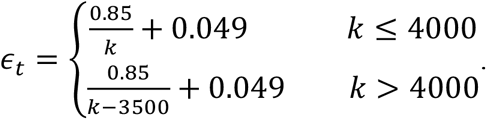. Typically, ∈ _*t*_ is set to decrease as *k* increases, thus balancing more exploration of random actions in the beginning and exploitation of the greedy actions in future iterations. We additionally defined *ϵ*_*t*_ to explore at a higher rate when *k* > 4000, to test for convergence. We initialize the Q-values to some constant (*C*), i.e., *Q*_*k*_(*x, t, a*) = *C*, ∀(*x, t, a*), for *k* = 0. We then iterate over *k* and all decision epochs within each iteration to update *Q*_*k+1*_(*x, t, a*) using immediate rewards *R*_*a*_(*x, x*′) returned by the simulator at every decision epoch *t* after simulating action *a*. The algorithm is known to converge to near-optimal solutions when *k* becomes a sufficiently large value [36]. That is, the optimal action (*say a*^*∗*^(*x, t*)) to be taken at time “*t*” when the system is in state “*x*” is defined as 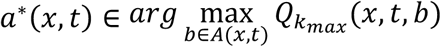. The schematic of the above iterative training process and summary of the above training steps of the algorithm are shown in Appendix section 5 (Figure A3 and Table A4 respectively).

The optimal policy ***d*** = [*d*_*0*_, *d*_*1*_, … *d*_*T*_] would then be identified by also using PATH 2.0 simulation along with trained Q-values 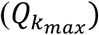. Specifically, we set *d*_*0*_ = *a*^*∗*^(*x*_*0*_, 0), where *x*_*0*_ =epidemic state in year 2015, simulate *a*^*∗*^, and say, the epidemic transitions to state *x*_*1*_ in 2020, then set *d*_*1*_ = *a*^*∗*^(*x*_*1*_, 1), simulate that action, and continue this iterative process until *T*. As the simulation is stochastic, we repeat this process 100 times to generate an uncertainty range.

#### Simulator-Progression and Transmission of HIV (PATH 2.0) model

PATH 2.0 is an agent-based stochastic simulation model that individually tracks HIV-infected persons by simulating HIV disease progression through a health-state transition model and sexual transmissions of HIV through a novel dynamic transmission model. PATH 2.0 is calibrated to be representative of the US HIV epidemic and has been validated to accurately simulate the epidemic for the years 2010 through 2015. Details of the model, its validation, and its adoption to inform HIV-related decisions in the US are presented elsewhere [26, 38].

We used the PATH 2.0 model to simulate an *action, a*(*x, t*), selected by the Q-learning algorithm at a decision epoch *t* given system state *x*, simulate *state transitions*, and calculate the corresponding *immediate reward R*_*a*_(*x, x*′) by assigning QALYs (*L*_*t*_) and costs (*C*_*t*_) to every person in the simulation. Specifically, for the selected proxy action (*a* = [*a*_*unaware,i*_, *a*_*ART,i*_; ∀*i*]), we estimate diagnostic and retention-in-care rates (Appendix section 2), and use PATH 2.0 to simulate diagnosis, care-and-treatment, and based on the care status of an infected person, simulate transmissions to their susceptible partners, thus transitioning the epidemic to a new state *x*^*r*^. We assign a QALY of 1 per year if the person is healthy, between 0 and 1 if HIV-infected (varying based on disease stage), and 0 if deceased [26, 38]. Based on estimated diagnostic and retention-in-care rates and the effectiveness of the intervention programs, we estimate the number of persons intervened for each program and corresponding costs, which are discussed in more detail in Appendix sections 2 and 4, respectively. Briefly, in the estimation of testing and retention-in-care costs, we made the following assumptions based on data from intervention programs [9, 39, 45]. HIV testing programs can be conducted in clinical or non-clinical settings, each having its own fixed and variable costs [39, 9]. Some people get tested voluntarily and incur only the cost of testing, while some get tested as a result of an outreach intervention and thus incur additional outreach costs [9], which we modeled as a non-linear function of the number of people outreached [15, 46]. In accordance with current CDC recommendations, we assumed only persons with high-risk are recommended for regular testing and intervened through outreach programs and that 6% of heterosexual females, 10% of heterosexual males, and all MSM are high-risk populations [47-49]. We assumed a non-linear variable cost function for retention-in-care to model the additional support programs needed to retain a larger proportion of people in care. Details of intervention costs are included in Appendix section 4.

In summary, we assumed that the first decision-making year is 2015 for the period 2016 to 2020, and decisions are made at every 5-year interval and solved for a decision sequence that optimally reaches close to zero new infections by 2070, i.e., in the Q-learning algorithm *t* ∈ {2015, 2020, …, 2070}. We used 2015 as the initial year as per the latest data available at the time of model development [26, 38]. However, the time-step of the simulation is monthly. Every iteration (*k*) of the Q-learning algorithm consists of simulating the PATH 2.0 model from 2015 to 2070 in monthly time-steps within a feedback and control loop to update Q-values and determine actions to be simulated every 5-years. Repeating this process for a large number of iterations, the optimizer learns to pick a better action at each iteration to eventually *converge* to an optimal *policy*. The model is coded in NetLogo 6.0.2 software [50].

#### Evaluating convergence of Q-learning algorithm to optimal policy

An algorithm has converged if it has reached a local optima through the iterative search process, i.e., successfully solved for an optimal combination of testing and retention-in-care rates. If the number of iterations is not sufficiently large, there is a risk that the algorithm is terminated before convergence. The ideal number is typically determined through experimentation. Further, there could be multiple local optima, i.e., multiple policies could yield similar total rewards, and because of the stochastic nature of the epidemic system, the optimal policy could be a range rather than a point estimate. Therefore, we ran the model for varying number of iterations, 2000, 3000, 4000, and 5000, and compared the corresponding total rewards (Appendix section 6, Figures A4 to A13), to ensure convergence and obtain the uncertainty range in optimal policies. The relative difference in the average costs and QALYs between the varying iterations were at most 2% in each cost function evaluated (see cost functions in Uncertainty Analysis section 3), suggesting convergence. The corresponding optimal policies differed slightly, more so in future years than earlier years, suggesting stochastic uncertainty as the model projects further into the future. Therefore, in Results, we present the range of optimal policies across these iterations as the uncertainty range.

## 3. Uncertainty analysis

We modeled two types of uncertainty:

1. The inherent stochasticity in the epidemic system is modeled through: a) the use of PATH 2.0, which is a stochastic simulation model where input parameters are drawn from probability distributions and events simulated using stochastic functions; b) the use of MDP with Q-learning, which is a stochastic control optimization method and; c) the use of varying numbers of MDP iterations (2000 to 5000), and simulating the optimal policy from each iteration a 100 times to generate the average values for output metrics.
2. The uncertainty in intervention costs is modeled by using three different cost functions, each with varying assumptions for the following four unit-costs a) the fixed cost per clinic for a retention-in-care program, b) the variable cost per person for a retention-in-care outreach program, c) the marginal increase in variable cost for a retention-in-care outreach program, and d) the marginal increase in variable cost for a testing outreach program [49]. These four parameters were chosen as they are related to support programs, which tend to have more variability as the type of support needed varies by individual-needs. As the model attempts to find the optimal balance in testing and retention-in-care rates, we selected a median and alternating bounds of the above four unit cost range to generate the overall range in optimal policy from uncertainty in costs. Therefore, we have three cost-functions as follows:

**Median (Median Testing and Retention-in-care Costs):** Uses the median values for all four parameters.

**LTHR (Low Testing High Retention in Care Costs):** Uses the lowest value for the testing costs and the highest value for the retention-in-care costs.

**HTLR (High Testing Low Retention in Care Costs):** Uses the highest values for the testing costs and the lowest values for the retention-in-care costs.

In summary, for each cost-function assumption, we trained the Q-learning algorithm with multiple stopping conditions (2000, 3000, 4000, and 5000 iterations). Using the trained Q-values 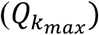, for each cost function and stopping condition pair, we simulated 100 runs and extracted the average values (over the 100 runs) of the optimal policy and corresponding impacts generated, specifically, values for the testing rate, retention-in-care rate, proportion of people with HIV (PWH) aware of their infection, proportion of PWH on ART, number of new infections, number of PWH, and incremental total costs, which are useful metrics from a public health perspective.

## 4. Results

For the end of 2015, the PATH 2.0 simulation model estimated an annual testing rate of 0.26 for high-risk heterosexuals and 0.4 for MSM, i.e., an average time from infection to diagnosis of 3.8 years for heterosexuals and 2.5 years for MSM. These results closely match the CDC estimates for the time from infection to diagnosis in 2015 [6], estimated using data from the NHSS [7]. For the end of 2015, the model estimated an annual retention-in-care rate of 86% for heterosexuals and 91% for MSM. Figure 1a (and 1b) presents the optimal policy for the period 2016 to 2070, specifically the optimal combination of testing (bottom) and retention-in-care (top) rates over time for heterosexuals (and MSM). Figure 1c (and 1d) shows the corresponding proportions aware (top) and on ART (bottom) for heterosexuals (and MSM). The figures present the uncertainty range (shaded bands) for each of the three cost function assumptions (Median: blue bands, LTHR: red bands, HTLR: green bands).

**Figure 1.**
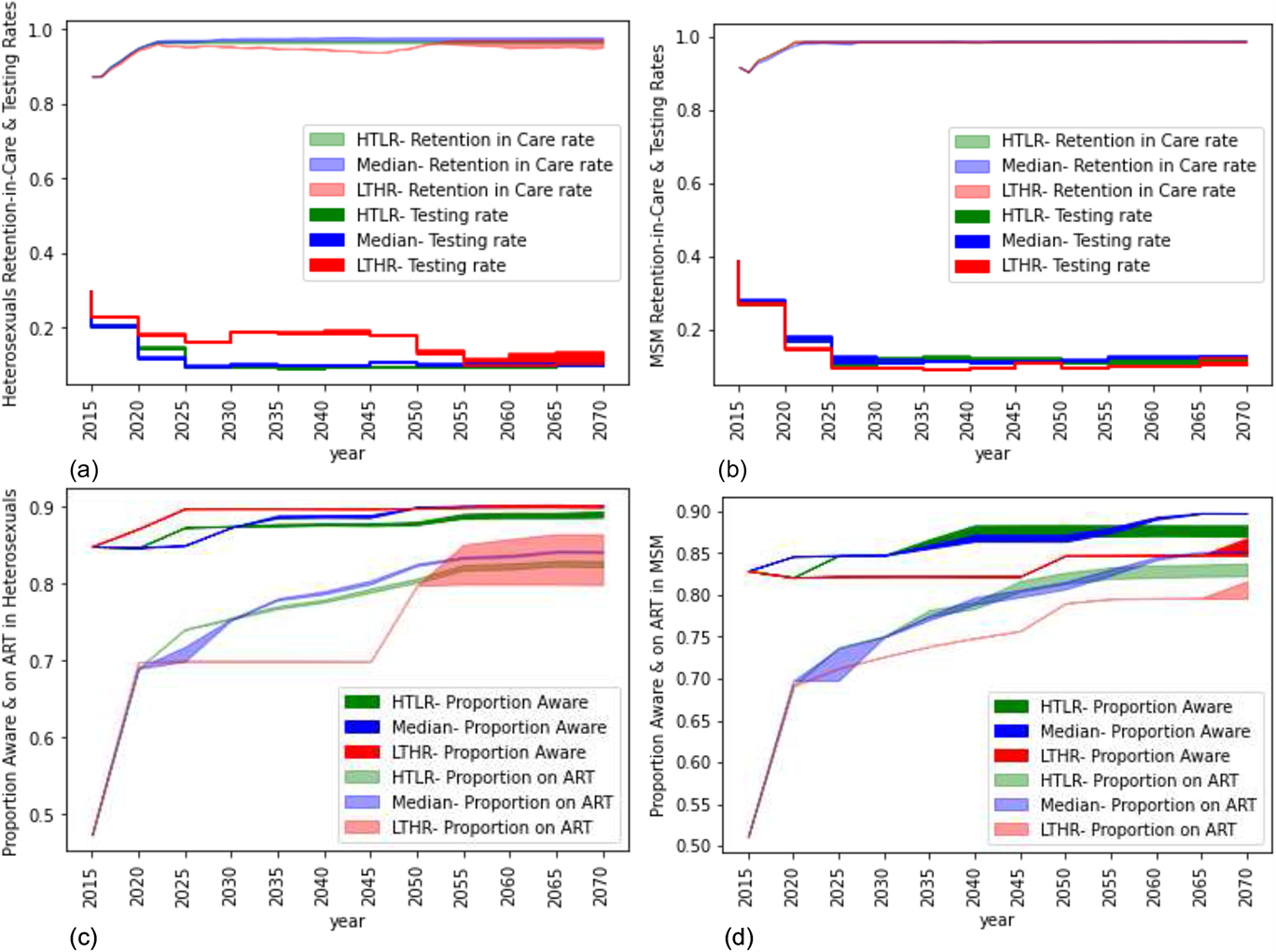
Optimal combinations of testing (top lines) and retention-in-care (bottom lines) rates for heterosexuals (1a) and MSM (1b) and corresponding proportion aware (top lines) and proportion on ART (bottom lines) for heterosexuals (1c) and MSM (1d) for three cost functions of HTLR (green), Median (Blue), and LTHR (Red). Results are an average of over 100 simulation runs of the optimal policy. The shaded region is the uncertainty around the optimal policy generated by the reinforcement learning algorithm.

For the period 2016 to 2020, under all three cost function assumptions, the model suggests a testing rate of 0.2 for high-risk heterosexuals (Figure 1a) and 0.3 for MSM (Figure 1b), equivalent to testing once every 5 and 3.5 years, respectively. Over the period 2016 to 2020, under all three cost function assumptions, the model suggests aggressive retention-in-care programs to gradually increase annual retention-rates from 86% to 94% for HETs (Figure 1a) and from 91% to 96% for MSM (Figure 1b). The uncertainty bands for this period, under all three cost function assumptions, for both testing and retention-in-care rates are narrow, suggesting a high chance that the algorithm has converged. Achieving the above testing and retention-in-care rates corresponded to about 85% of all heterosexual PWH and 82% of all MSM PWH being aware of their infection by the end of 2020, and about 70% of all heterosexual PWH and 70% of all MSM PWH on ART by the end of 2020 (Figure 1c and 1d).

Implementing the combination of testing and retention-in-care rates for the period 2016 to 2020 generated a 50% reduction in annual new infections among heterosexuals (from 9000 in 2016 to 4500 by the end of 2020) and a 42% reduction in annual new infections among MSM (from 26,000 in 2016 to 15,000 by the end of 2020) which is a significant reduction compared to trends over the previous 5-year period (Figure 2a and 2b). There was a modest decline in the number of heterosexual PWH, breaking the previous trend of growth (Figure 2c). There was modest-growth in the number of MSM PWH, which is a shift from the previous high-growth rate but suggests that the number of PWH will continue to increase for a short period before declining (Figure 2d). The annual cost of HIV increased by about 22% over this period, suggesting a high initial investment to achieve the above reduction in new infections (Figure 3).

**Figure 2.**
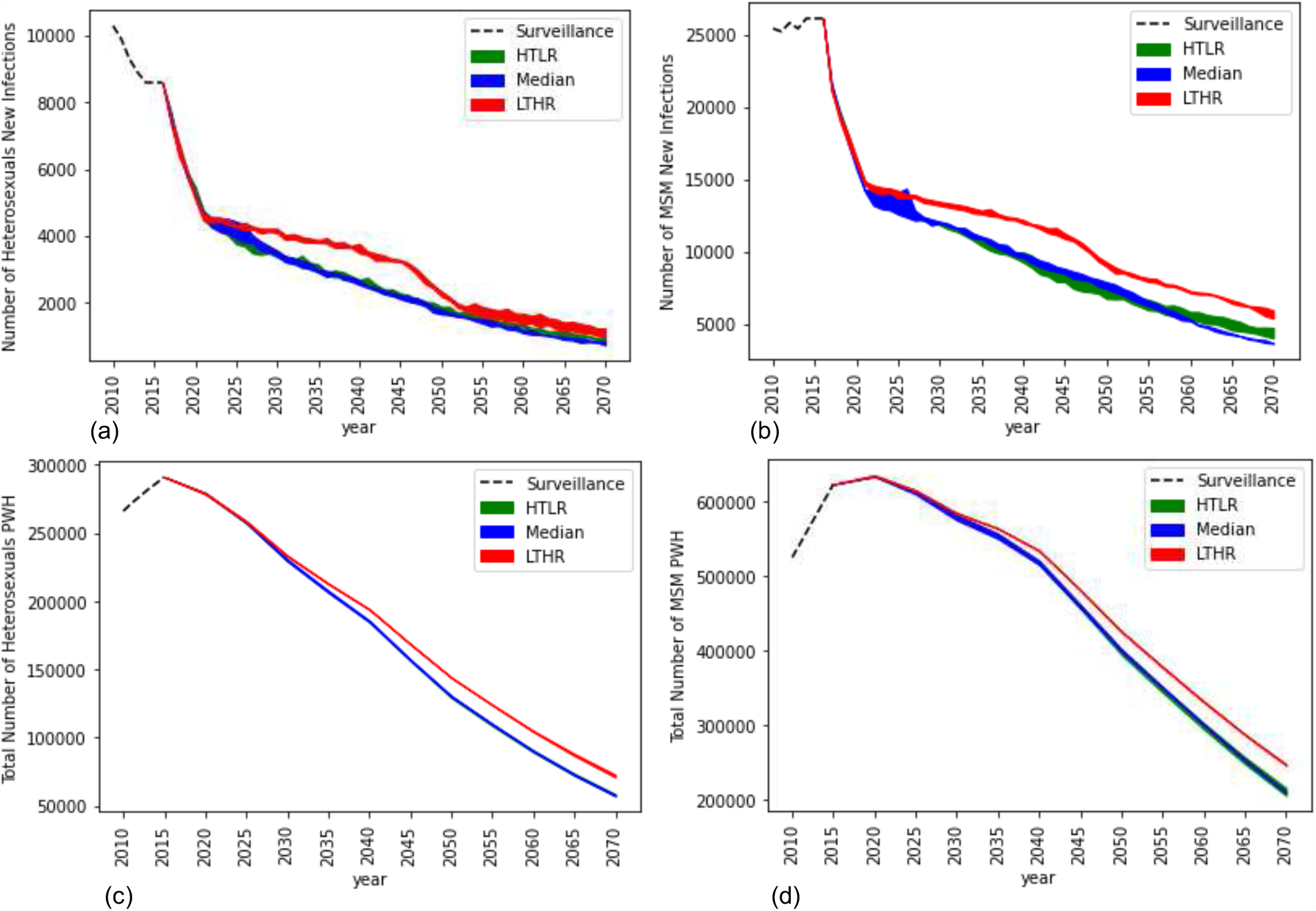
The number of new infections for heterosexuals (1a) and MSM (1b) and the number of people living with HIV for heterosexuals (1c) and MSM (1d) for the HTLR (green), Median (Blue), LTHR (Red) cost function assumptions. Results are an average of over 100 simulation runs of the optimal policy. The shaded region indicates the uncertainty range in the optimal policy.

**Figure 3.**
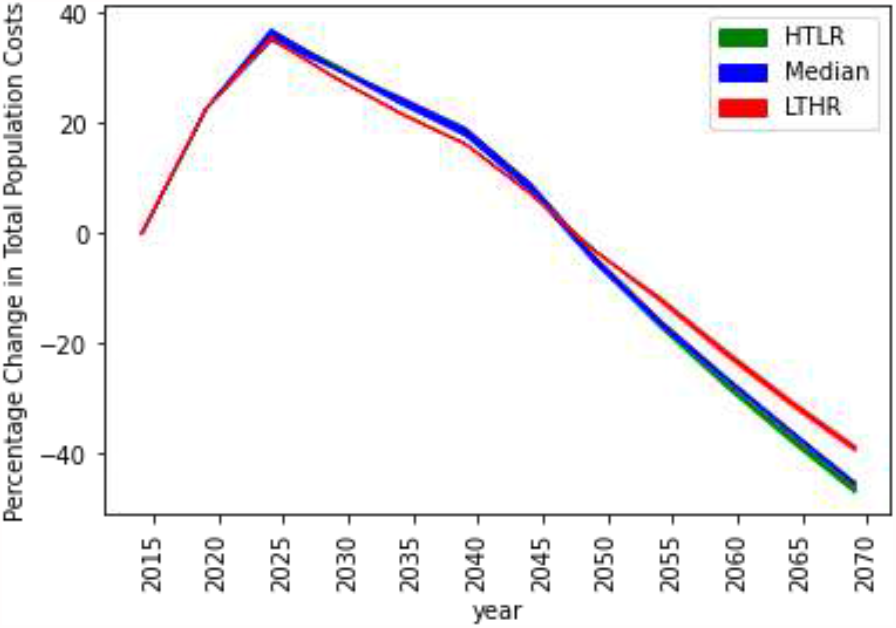
Changes in total population costs corresponding to the optimal policy under the three cost function assumptions, HTLR (green), Median (Blue), LTHR (Red).

For the period 2021 to 2025, compared to the previous 5-year period, the model suggests relaxing the frequency of testing while modestly increasing and maintaining a high retention-in-care rate for both heterosexuals and MSM and under all three cost function assumptions. Specifically, for heterosexuals, the model suggests testing rates of 0.14, 0.11, and 0.18 under the HTLR, Median, and LTHR cost assumptions, equivalent to a testing frequency of every 7, 9, and 5.5 years, respectively (Figure 1a). For MSM, the model suggests a testing rate of around 0.15 under all three cost function assumptions, equivalent to a testing frequency of every 6.5 years (Figure 1b). The model suggests scaling-up retention-in-care programs to increase annual retention-rates from 94% to 96% for heterosexuals (Figure 1a), and from 96% to 98% for MSM (Figure 1b) over the period 2021 to 2025. The reduction in new infections was modest compared to the previous 5-year period for both heterosexuals (Figure 2a) and MSM (Figure 2b), but the number of PWH declined at a faster rate compared to the previous 5-year period for heterosexuals (Figure 2c), and for the first time there was a reduction in the number of MSM PWH (Figure 2d).

For the period 2026 to 2030, the testing rate reduced to 0.1 (test once every 10 years or less) under Median and HTLR cost functions in heterosexuals (Figure 1a) and under all cost functions for MSM (Figure 1b) and continued to remain at those values for the remaining duration of the simulation (up to 2070). Under the LTHR cost function for heterosexuals, the testing rate continued to remain at 0.18 (test once every 5.5 years) until 2050 and then reduced to 0.1 for the remaining duration of the simulation. The corresponding proportion aware and on ART gradually increased to 80% (Figure 1c and 1d), and the corresponding number of new infections (Figure 2a and 2b) and PWH (Figure 2c and 2d) gradually decreased over the period 2026 to 2070 for both heterosexuals and MSM. For heterosexuals (Figure 2a), under the three cost function assumptions, the number of new infections reduced to about 3200 to 4000 cases per year by 2030 (53% to 62% reduction compared to 2015) and further reduced to 750 to 1200 cases per year by 2070 (86% to 91% reduction compared to 2015). For MSM (Figure 2c), under the three cost function assumptions, the number of new infections reduced to about 11,000 to 14,000 cases per year by 2030 (46% to 58% reduction compared to 2015), and further reduced to 3500 to 6000 cases per year by 2070 (80% to 86% reduction compared to 2015).

Comparing across the cost function assumptions, for heterosexuals, the optimal rates were generally intuitive, with the highest testing and lowest retention-in-care in LTHR and lowest testing and highest retention-in-care rates in HTLR, though the differences in retention-care rates were modest (Figure 1a). For MSM, however, the optimal annual retention-in-care rates were similar in all three cost functions, and the results were counter-intuitive for testing rate as the model suggested a slightly lower testing rate in LTHR compared to Median and HTLR. That is, some of the testing resources were shifted to retention-in-care programs to offset its higher costs while maintaining annual retention-rates at the level of Median and HTLR (Figure 1b). And as a result, the proportion of MSM on ART in LHTR, though lower than in Median and HTLR, was higher than that of heterosexuals on ART in LHTR (Figure 1d), which suggests the higher significance of retention-in-care programs to ensure sustained care-and-treatment, compared to testing. The optimality of this counter-intuitive strategy in MSM was evaluated by a counterfactual simulation run using the optimal LTHR strategy of heterosexuals for both heterosexuals and MSM. The number of new infections, PWH, and costs were higher in the counterfactual simulation, confirming the optimality of the strategy. Details of this run and the results are presented in Appendix section 7.

## 5. Discussion and conclusion

This paper proposes a methodology for phased-decision-making, which is typical in public health for epidemic control. We modeled the question of ‘how to optimally reach HIV elimination in the US’ as a sequential decision-making problem by formulating it as an MDP and solving it using a Q-learning algorithm. Compared to the most commonly used approach of explicitly evaluating a few pre-selected scenarios, our approach enables selecting an optimal from all possible choices (36^11^ possible choices) through probabilistic projections of the decisions and the epidemic. In identifying the optimal sequence of combinations of testing and retention-in-care rates, we took a societal perspective to evaluate costs and QALYs. Though the stochastic dynamic sequential decision-making models are very attractive for evaluations of phased decision-making, they are computationally expensive to solve as the state space and action space of epidemic control problems are typically large, giving rise to issues of non-convergence and thus limiting its applicability. In this study, we reformulated the action space by taking indirect metrics that significantly reduced the size of the action space, thus leading to a successful application. Though applied to HIV, the proposed approach can be used for other infectious diseases as typically testing and treatment are key methods to control spread.

Our analysis is subject to limitations. We constrained the maximum intervention scale-up to one time and two times the maximum scale-up achieved in the past five years for proportion aware and proportion on ART, respectively, to balance feasibility and aggressive scale-ups. We also did not consider the availability of new interventions in the future, such as vaccines. We did not consider preventive interventions such as pre-exposure prophylaxis or changes in sexual behaviors in future generations. We did not evaluate interventions specific to people who inject drugs. We did not consider changes in demographics over time. Limitations to any model-based analysis, such as any drawbacks from the quality of data and unavailability of data leading to parameter estimations rather than the use of actual data, also apply to the PATH 2.0 simulation model and are discussed elsewhere [26]. Data on costs of support services and outreach are limited, and thus, we made assumptions on its variability to evaluate its sensitivity to optimal decisions.

Despite these limitations, we believe the approach used in this paper in evaluating phased-decisions related to the two most effective interventions is very timely in light of the ‘Ending the HIV Epidemic’ federal plan for an HIV elimination objective [2]. The optimal policies generally suggest more frequent testing for the first 10 years and less frequently after as the number of new infections decreases and the proportion aware increases. It suggests more frequent testing for MSM than heterosexuals for the first 10 years, which supports the risk-based testing proposed under current CDC guidelines [51], and similar testing rates for both risk groups thereafter. It suggests implementing retention-in-care programs to gradually increase annual retention-in-care rates to 95% within the first 10 years and maintain it at that rate thereafter. Generally, the model suggested aiming for a higher retention-in-care rate than the testing rate, suggesting prioritization of spending on retention-in-care programs. Optimal decisions were mostly robust to uncertainty in costs, under the range assumed, except for MSM under LTHR. In this scenario, the model suggested taking advantage of the lower testing costs and maintaining a higher testing rate for a longer duration than in the Median and HTLR cost functions. While testing and retention-in-care rates help inform intervention programs, the corresponding targets for proportions aware and on-ART serve as control measures, i.e., help direct resources to relevant support programs and risk groups in cases where actual proportions (that are tracked through NHSS) fall short of the target proportions. Proportions aware and on-ART are leading indicators in the EHE, and thus, actual proportions are regularly tracked as part of the NHSS.

The federal plan for ‘Ending the HIV Epidemic’ aims for a 75% reduction in annual new infections by 2025 (to 9,300 new cases per year) and a 90% reduction by 2030 (to 3,000 new cases per year) [2, 3]. Our results indicate that testing and retention-in-care programs alone would be insufficient to reach this goal optimally. The optimal strategy would achieve a significant reduction in annual incidence by 2030, reducing to about 14,200 to 18,000 annual new infections, which is a 50% to 60% reduction from 2015, but then gradually decrease at a slower rate thereafter, reaching about 4,250 to 7,200 annual new infections by 2070, which is an 86% to 91% reduction from 2015. To further accelerate the reduction in new infections, other novel interventions that are recently emerging, such as pre-exposure prophylaxis for HIV-negative individuals and more targeted testing through identification of transmission clusters, should be explored [2].

## Data Availability

This model is a simulation-based optimization model. Data for the simulation and optimization model are gathered from different sources indicated in the manuscript.

## Appendix A reinforcement learning model to inform optimal decision paths for HIV elimination

### 1. Depicting MDP as a decision tree

The sequential decision-making problem for HIV epidemic control and elimination can be visualized as a decision-tree. Figure A1 presents the decision tree for a simple 2-state system with *n* action choices, with the epidemic state in 2015 being the start node, actions being the decision nodes, epidemic states at 5-year intervals being the chance nodes, the possible epidemic states in 2070 being the end nodes, and a policy being a path in a decision-tree. The problem is to identify, from among all possible paths of a decision-tree, the one with the maximum total reward. The decision tree starts in the initial state *S*_*O*_ and corresponds to the epidemic state in the year 2015 (*t*_*O*_). Each square is a decision node at which there are multiple actions (stems branching out from a node) to choose from ([*a*_*l*_, *a*_*2*_, … *a*_*n*_] in Figure A1). An action taken in any year *t*_*i*_ transitions the epidemic to a different state in year *t*_*i+S*_, with some uncertainty (oval) in the actual state it transitions to, denoted as probabilities *p*_*a*_(*j, k*) for transitioning to state *S*_*k*_ from state *S*_*j*_ when action *a* is taken. Thus, the decision tree branches out from year the 2015 to 2070, with decision-making occurring at every 5-year intervals. The “total reward” is the output at the end of each branch in the year 2070. However, the problem cannot be solved using decision trees because it is impractical to evaluate all possible paths exhaustively. In this simple 2-state *n*-actions example, that would be 2*n* paths if evaluating up to time *t*_*l*_, (2*n*)^*2n*^ paths if evaluating up to time *t*_*2*_, thus growing exponentially over the time-periods evaluated.

The HIV model introduced in this paper is significantly more extensive and has 16,501-states 36-actions evaluated for 11 time-periods. Hence, we formulated the problem as a Markov decision process (MDP), solved using optimization modeling that efficiently uses mathematical concepts to search through the solution space. MDP can also be solved by dynamic programming or reformulating as a linear programming optimization model; however, these methods also require the knowledge of the transition probability matrices (*P*_*a*_) for each action *a*, square matrices with dimensions equal to the number of states (*p*_*a*_(*j, k*) are elements of *P*_*a*_), which is infeasible for HIV [1]. Therefore, to overcome these challenges, we use Q-learning with a stochastic dynamic simulation model to directly simulate the state transitions (replacing *P*_*a*_) and estimate rewards.

**Figure A1:**
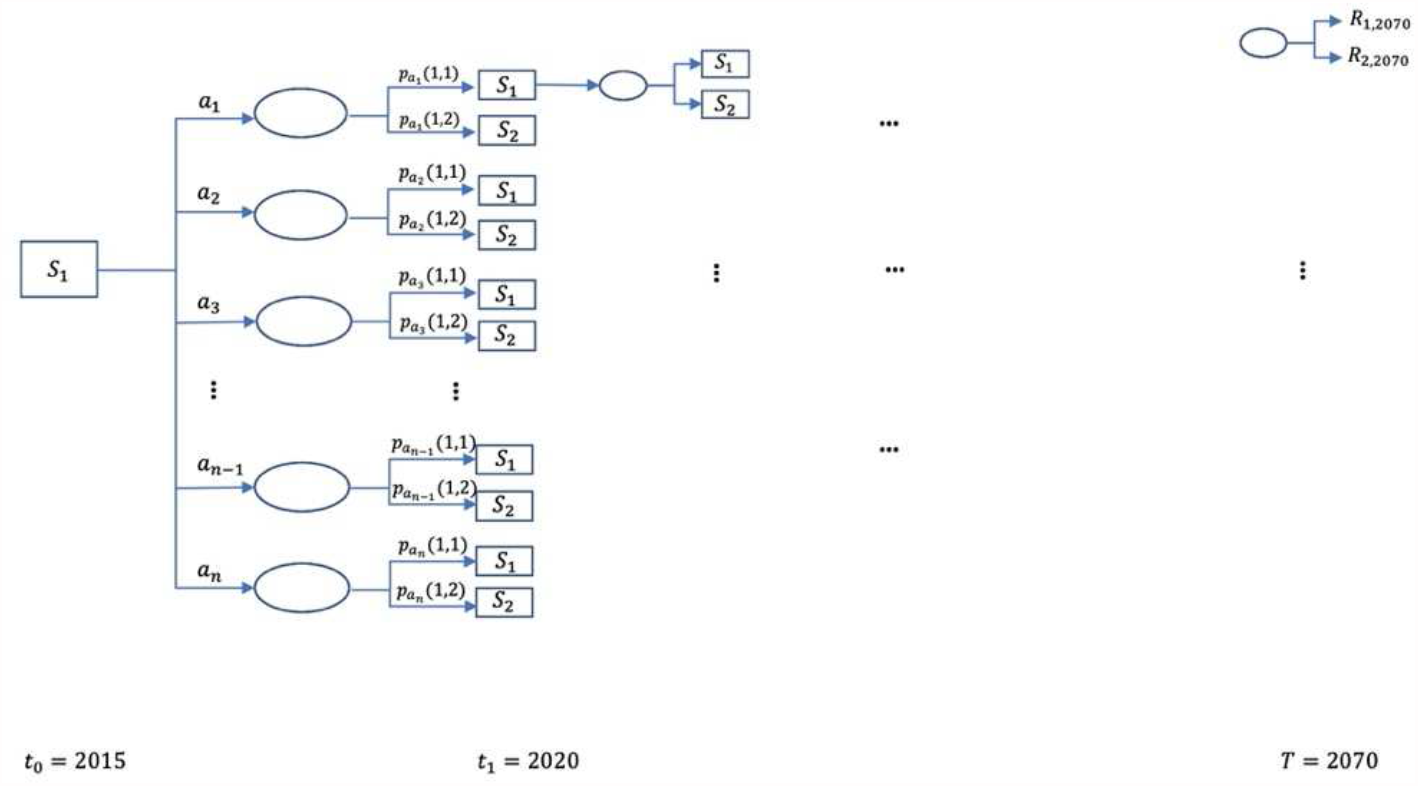
Decision tree representation of the Markov decision process problem (MDP); the objective of MDP is to find the path with an optimal trade-off in costs and benefits.

### 2. Estimating diagnostic and retention-in-care rates for a given action and simulating the action in PATH 2.0 model

As discussed in the main manuscript, we formulated an action as a combination of the percentage decrement in proportion unaware and the percentage increment in proportion on ART; each varying by transmission risk-group (*a*_*unaware,i*_, *a*_*ART,i*_, ∀*i* ∈ {*HETs, MSM*}). Corresponding to every action, we estimated the diagnostic rate (*δ*_*t*_) and retention-in-care rate (1 − *ρ*_*t*_), and used the rates for simulating the action in the Progression and Transmission of HIV (PATH 2.0) model [2], including estimating the number of persons successfully intervened (newly diagnosed and retained-in-care) and the corresponding intervention costs. The estimation method includes a combination of expressing the system as a differential equations model and utilizing the PATH 2.0 model. In this section, we present the method of estimating *δ*_*t*_ and 1 − *ρ*_*t*_ using *a*_*unaware,i*_ and *a*_*ART,i*_, the corresponding number of persons successfully intervened (newly diagnosed and retained-in-care), and simulating the action in PATH 2.0. In addition, we present the estimation of the intervention costs of that action in Appendix section 4.

We can express the disease incidence and transition along the stages of care-continuum of HIV-infected persons as a set of differential equations or a compartmental model, with each disease and care continuum stage representing a compartment (Figure A2).**Error! Reference source not found**. We have four compartments, susceptible (S), infected and unaware (U), infected and aware but not in care (NC), and infected in care and on ART (ART).

**Figure A2:**
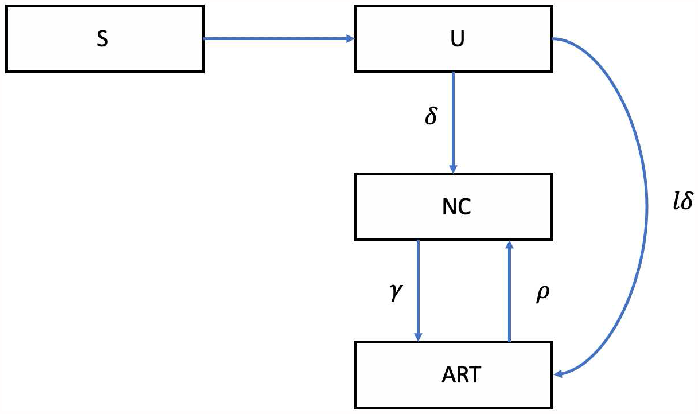
Flow diagram for disease incidence and transitioning along the stages of care-continuum HIV infected persons. S: susceptible, U: infected and unaware, NC: infected and aware but not in care, ART: infected, in care and on ART, δ: diagnostic rate, γ: rate of entering care and treatment among those not in care, and ρ: rate of dropping-out of care, and lδ: rate of diagnosis and linked to care.

Let,

*I*_*t*_ be the number of infected persons at time *t*,

*i*_*t*_ be the number of new infections at time *t*,

*p*_*t,s*_ be the proportion of infected persons in stage s ∈ {U, NC, *ART*}, such that *p*_*t,s*_*I*_*t*_, would be the number of infected people in stage s at time *t*,

*δ*_*t*_ be the diagnostic rate at time *t*,

*l*_*t*_ be the proportion of persons linking to care and initiating treatment upon diagnosis, among those diagnosed at time *t*,

*γ*_*t*_ be the rate of entering care and treatment among those not in care at time *t*,

*ρ*_*t*_ be the rate of dropping-out of care and treatment at time *t*, and

*m*_*t*_ be the number of new deaths at time *t*.

Then, at a sufficiently small incremental time-step *t* + 1 (we use monthly increments), we can write the equations for the number of people in each stage by formulating as a system of differential equations,

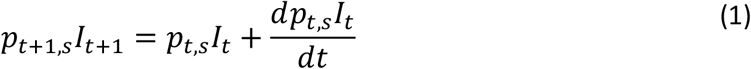

where, 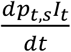 is the rate of the change in *p*_*t,s*_*I*_*t*_, i.e., the change in the number of infected persons in stage s at *t*.

Specifically, for each stage s ∈ {U, NC, C} we can apply (1) and write,

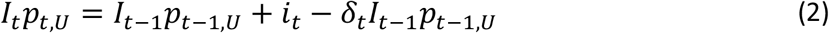

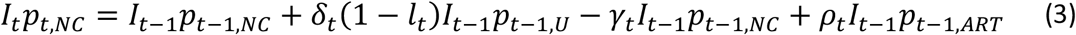

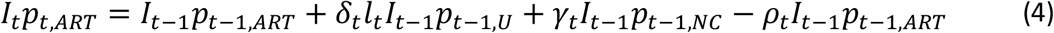

Additionally, as *p*_*t,s*_ is the ‘proportion’ of infected people in stage s, summing over all disease stages should add to 1, or

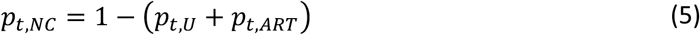

Further, the number of infected people at *t* (*I*_*t*_) would be the number infected at *t* − 1 plus new infections (*i*_*t*_) minus deaths (*m*_*t*_), i.e.,

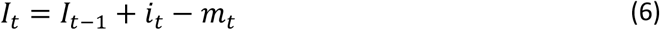

Note, these equations are applied for each risk group (heterosexuals, and MSM) separately, but we do not indicate the risk group in subscripts for clarity of notations.

#### Estimate diagnostic rate (*δ*_*t*_) and the corresponding number of persons successfully intervened (newly diagnosed)

We can write the monthly change in proportion unaware as −(*p*_*t − 1,U*_ − *p*_*t,U*_)*=* â _*unaware,t*_, which is the selected proxy action choice for the change in proportion unaware at decision epoch T divided by 60 months (*i*. e., 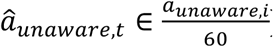), as our decision-making interval is every five years. Note that, â _*unaware,t*_ is specific to risk-group *i* but we drop subscript *i* for clarity of notation. We calculate monthly estimates as the time-step in the PATH 2.0 simulation model is monthly. We assume the changes in proportion unaware are achieved uniformly over the 60-month period, and thus, the estimated diagnostic rate would be representative of linearly scaling up HIV-testing interventions over this interval. Then, using equation (6), we can express diagnostic rate by rewriting (2) as

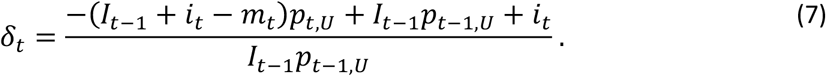

By substituting *p*_*t,U*_ = *p*_*t−1,U*_ + â _*unaware,t*_ in equation (7) we can write

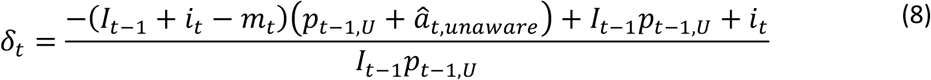

and the corresponding number of persons to diagnose as

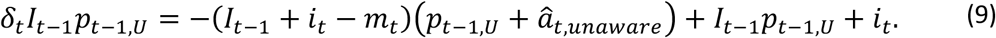

#### Estimate retention-in-care rate and the number of persons successfully intervened (retained-in-care)

We can write the monthly change in proportion on ART as (*p*_*t,ART*_ − *p*_*t − 1,ART*_) = â _*ART,t*_, which is the selected proxy action choice for the change in proportion on ART at decision epoch T divided by 60 months (*i*. e., 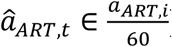), as our decision-making interval is every five years. Note that, â _*ART,t*_ is specific to risk-group *i* but we drop subscript *i* for clarity of notation. We assume the changes in proportion on ART are achieved uniformly over the 60 month period, and thus, the estimated retention-in-care rates would be representative of linearly scaling up retention-in-care interventions over this interval. We set *p*_*t,ART*_ = *p*_*t − 1,ART*_ + â _*ART,t*_ in equation (4) and rewrite it to express the rate of dropping-out as

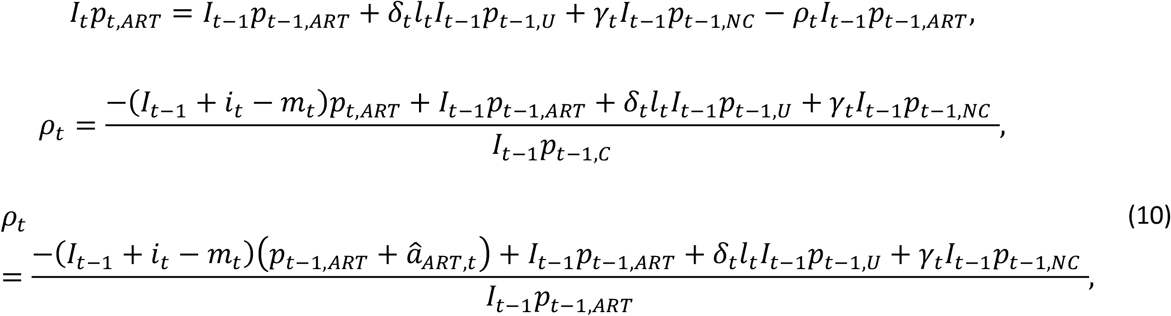

and estimate the corresponding number of persons dropping-out of care as

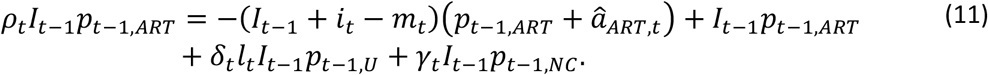

Following from above, we estimate the number of persons retained in care as the number of persons not dropping-out of care, i.e., (1 − *ρ*_*t*_)*I*_*t − 1*_*p*_*t − 1,ART*_.

#### Simulating the action in PATH 2.0

Simulating any given action *a*_*unaware,i*_, *a*_*ART,i*_, ∀*i* ∈ {*HETs, MSM*} over the 5-year interval (in monthly time-steps) involves iteratively simulating PATH every-month for estimating the terms on the right-hand side of equations (8) and (10), solving for diagnostic rate (*δ*_*t*_) and retention-in-care rate (1 − *ρ*_*t*_) for that month using (8) and (10), estimating the numbers to diagnose and retain-in-care using (9) and (11), and using that in PATH to simulate diagnosis and care events for that month. Details of the PATH 2.0 model along with the estimation of the parameters in (8) and (10) and simulating care and diagnosis events are discussed elsewhere [2], here we only give a brief description related to the estimation of parameters in (8) and (10). PATH 2.0 model was initialized to be representative of the HIV infected population in the US in 2006, using data from the US National HIV Surveillance System (NHSS), and simulated to 2015. It keeps track of *I*_*t*_ and *p*_*t,s*_ over time. It estimates new infections (*i*_*t*_) by modeling transmissions at the individual-level for every susceptible-infected partnership, modeling it as a function of stage s ∈ {U, NC, *ART*} of the infected person at time *t* − 1. Thus, the number of new infections is a function of *p*_*t − 1,U*_ and *p*_*t − 1,NC*_. It estimates the number of deaths (m_*t*_) by simulating mortality at the individual-level for each person using stage-and age-specific rates of mortality. It uses annual data from NHSS for the proportion linking to care (*l*_*t*_) upon diagnosis and assume it will be maintained at the 2015 level for future years [3]. It uses re-entry rates (γ_*t*_) from studies in the literature [4].

### 3. Mathematical advantage of formulating action space as changes in proportions unaware and on ART instead of diagnostic and retention-in-care rates

As discussed in section 2, we formulate the action space (A) as a combination of changes in proportions unaware and on ART, i.e., A = {[*a*_*unaware,i*_, *a*_*ART,i*_, ∀*i* ∈ {*HETs, MSM*}]}, as proxy for diagnostic and retention-in-care rates {[*δ*_*i*_, 1 − *ρ*_*i*_, ∀*i* ∈ {*HETs, MSM*}]}. We prove here that the proxy metrics efficiently constrain the size of the action space, which would increase the chance of convergence of the Q-learning algorithm. It is also computationally efficient as it requires fewer evaluations of the simulation model. We also prove that formulating an action as [Δ*δ*_*i*_, 1 − Δ*ρ*_*i*_, ∀*i* ∈ {*HETs, MSM*}] also leads to a large action space, where Δ*δ*_*i*_ and 1 − Δ*ρ*_*i*_ are changes in *δ*_*i*_ and *ρ*_*i*_, respectively, over two consecutive decision epochs, as the proxies *a*_*unaware,i*_ and *a*_*ART,i*_ are also decrements or increments (of *μ*_*u,i*_ or *μ*_*v,i*_, respectively) over two consecutive decision epochs. We also prove that, corresponding to every combination of [â _*unaware,t*_, â _*ART,t*_] there is a unique combination of [*δ*_*t*_, 1 − *ρ*_*t*_], and thus solving for the optimal combination of the proxy metrics is equivalent to solving for the optimal diagnostic and retention-in-care rates. We prove these through Remarks 1 to 4.

Note that, [â _*unaware,t*_, â _*ART,t*_] and [*δ*_*t*_, 1 − *ρ*_*t*_] have *t* (time) subscripts as they are the monthly values defined in Appendix section 2 corresponding to a proxy action [*a*_*unaware,i*_, *a*_*ART,i*_, ∀*i*] and original action {[*δ*_*i*_, 1 − *ρ*_*i*_, ∀*i*]}, respectively. They do vary by risk-group but we drop the subscripts *i* for clarity of notations. Without loss of generality, we use [â _*unaware,t*_, â _*ART,t*_] and [*δ*_*t*_, 1 − *ρ*_*t*_] to prove our Remarks about [*a*_*unaware,i*_, *a*_*ART,i*_, ∀*i* ∈ {*HETs, MSM*}] and [*δ*_*i*_, 1 − *ρ*_*i*_, ∀*i* ∈ {*HETs, MSM*}], respectively.

#### Remark 1

Given the system state *x* at time *t* − 1, (*X*_*t − 1*_ = *x*), corresponding to every action, *a*_*unaware,i*_, there is a unique diagnostic rate, δ_*t*_, i.e., *f*: â _*unaware,t*_ → *δ*_*t*_ is a bijection function and corresponding to every action, *a*_*ART,i*_, there is a unique retention-in-care rate, 1 − *ρ*_*t*_, i.e., *g*: â _*ART,t*_ → 1 − *ρ*_*t*_ is a bijection function.

#### Proof

From equation (8) we have diagnostic rate as:

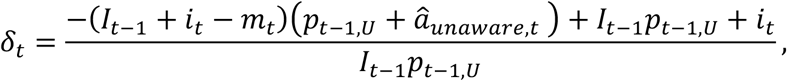

the only controllable unknown is â _*unaware,t*_ as all other parameters correspond to or are calculated using system state at time *t* − 1, as discussed in section 2. Therefore, *δ*_*t*_ is a linear function of â _*unaware,t*_, i.e., *f*: â _*unaware,t*_ → *δ*_*t*_ is a bijection function. This implies that, at any given system state at time *t* − 1 for every action â _*unaware,t*_, we can calculate a unique value for the diagnostic rate.

Similarly, from equation (10) we have drop-out rate as:

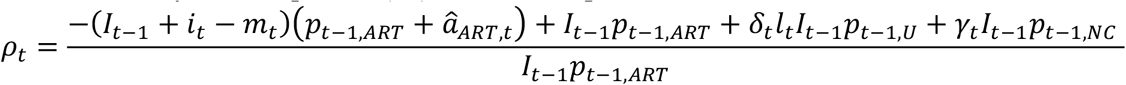

the only controllable unknown is â _*ART,t*_ as all other parameters correspond to or are calculated using system state at time *t* − 1, as discussed earlier in section 2. Therefore, *ρ*_*t*_ is a linear function of â _*ART,t*_, i.e., *g*: â _*ART,t*_ → *ρ*_*t*_ is a bijection function. This implies that, at any given system state at time *t* − 1, for every action *a*_*ART*_, we can calculate a unique value for the retention-in-care rate 1 − *ρ*_*t*_.

#### Remark 2

From a public health perspective, all actions that result in (*p*_*t − 1,U*_ − *p*_*t,U*_) < 0 or (*p*_*t,ART*_ − *p*_*t − 1,ART*_) < 0 are undesirable and should not be selected.

#### Proof

If the conditions are true, it would imply that a larger proportion of people with HIV are unaware of their infection and/or are not on treatment at time t compared to *t* − 1. Being unaware and not on treatment are associated with an increase in transmissions and mortalities. Thus, all actions that result in (*p*_*t − 1,U*_ − *p*_*t,U*_) < 0 or (*p*_*t,ART*_ − *p*_*t − 1,ART*_) < 0 are undesirable as they worsen the epidemic and should not be selected.

#### Remark 3

Setting action space as A = {[*a*_*unaware,i*_, *a*_*ART,i*_, ∀*i*]} instead of A = {[*δ*_*i*_, 1 − *ρ*_*i*_, ∀*i*]} efficiently controls the number of possible interventions and thus is more computationally efficient.

#### Proof

If action space A = {[*a*_*unaware*_, *a*_*ART*_]}:

As â _*unaware,t*_ = − (*p*_*t−1,U*_ − *p*_*t,U*_), we can directly select actions such that −(*p*_*t − 1,U*_ − *p*_*t, U*_) ≥0 (see Remark 2). As we use *a*_*unaware*_ to estimate diagnostic rate, it constrains diagnostic rates to only those that correspond to desirable outcomes. Thus, all testing intervention programs that are below the minimum diagnostic rates can be excluded. The selection of action is not dependent on any parameters of the system, except for the feasibility constraint p_*t,U*_ > 10%, as discussed in section 2.1 of the manuscript.

Similarly, as *a*_*ART*_ = −(*p*_*t − 1,ART*_ − *p*_*t,ART*_), we can directly select actions such that (*p*_*t,ART*_ − *p*_*t − 1,ART*_) ≥0 (see Remark 2), naturally constraining the selection of retention-in-care rates to those that would result in desirable outcomes. Thus, all intervention programs whose efficacy is below the minimum retention-in-care rates estimated here can be excluded. The selection of action is not dependent on any parameters of the system, except for the feasibility constraint *p*_*t,ART*_ < 90%, as discussed in the section 2.1.

Rearranging equation (7) for diagnostic rate from above, we can write

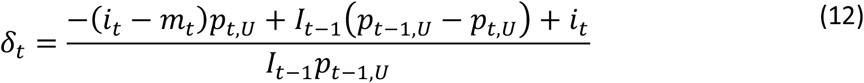

And further rearranging to write

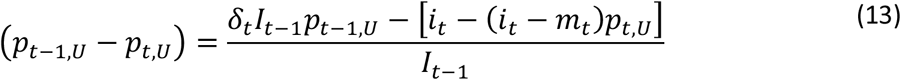

As *p*_*t,U*_ ≤ 1, if *i*_*t*_ > *m*_*t*_ then −(*i*_*t*_ − *m*_*t*_)*p*_*t, U*_< *i*_*t*_; and if *i*_*t*_ < *m*_*t*_ then −(*i*_*t*_ − *m*_*t*_)*p*_*t,U*_ > 0. Therefore, the following condition is always true [*i*_*t*_ − (*i*_*t*_ − *m*_*t*_)*p*_*t,U*_] > 0.

The above implies that, there are certain values of δ_*t*_ such that *δ*_*t*_*I*_*t − 1*_*p*_*t − 1*,_ ≥ [*i*_*t*_ − (*i*_*t*_ − *m*_*t*_)*p*_*t,U*_], which would yield (*p*_*t − 1, U*_ − *p*_*t,U*_) ≥ 0, and certain other values of *δ*_*t*_ such that *δ*_*t*_*I*_*t − 1*_*p*_*t − 1,U*_ < [*i*_*t*_ − (*i*_*t*_ − *m*_*t*_)*p*_*t,U*_], which would yield (*p*_*t − 1,U*_ − *p*_*t,U*_) < 0, which is an undesirable outcome from a public health perspective. As the values of δ_*t*_ that generate *δ*_*t*_*I*_*t − 1*_*p*_*t − 1*,_ ≥ [*i*_*t*_ − (*i*_*t*_ − *m*_*t*_)*p*_*t,U*_] is time-dependent on values of *I*_*t − 1*_, *p*_*t − 1,U*_, *i*_*t*_, *a*nd *m*_*t*_, a large set of values for *δ*_*t*_ should be evaluated as part of the action space. This is computationally expensive, and moreover inefficient, as many cases would result in an undesirable outcome.

Rearranging the equation for drop-out rate from equation (10), we can write

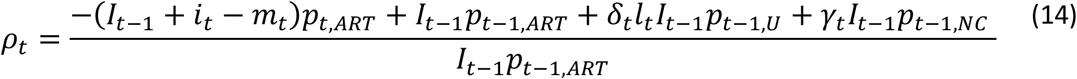

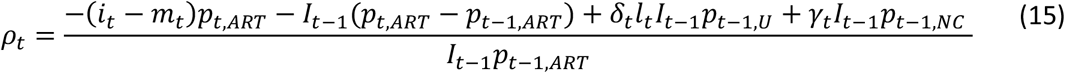

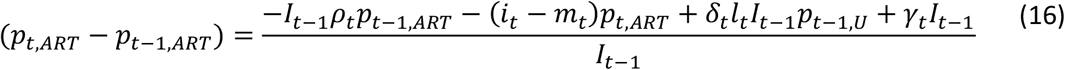

If *i*_*t*_ > *m*_*t*_ then (*i*_*t*_ − *m*_*t*_)*p*_*t,ART*_ > 0 and as +*δ*_*t*_*l*_*t*_*I*_*t − 1*_*p*_*t − 1*,_ + *γ*_*t*_*I*_*t − 1*_ > 0 and −*I*_*t − 1*_*ρ*_*t*_*p*_*t − 1,ART*_ < 0, there are certain combinations that can result (*p*_*t,ART*_ − *p*_*t − 1,ART*_) <0. Similarly, if *i*_*t*_ < *m*_*t*_ then (*i*_*t*_ − *m*_*t*_)*p*_*t,ART*_ < 0 and as +*δ*_*t*_*l*_*t*_*I*_*t − 1*_*p*_*t − 1,U*_ + *γ*_*t*_*I*_*t − 1*_ > 0 and −*I*_*t − 1*_*ρ*_*t*_*p*_*t − 1,ART*_ < 0, there are certain combinations that can result in (*p*_*t,ART*_ − *p*_*t − 1,ART*_) <0.

As the values of ρ_*t*_ that result in (*p*_*t,ART*_ − *p*_*t − 1,ART*_) <0 are time-dependent on values of *I*_*t − 1*_, *p*_*t − 1,ART*_, *i*_*t*_, and *m*_*t*_ a large set of values for *ρ*_*t*_ and thus 1 − *ρ*_*t*_ should be part of the action space, which is again computationally expensive and inefficient as many cases would result in undesirable outcomes.

#### Remark 4

Setting action space as A = {[*a*_*unaware,i*_, *a*_*ART,i*_, ∀*i*]} instead of A = {[Δ*δ*_*i*_, 1 − Δ*ρ*_*i*_, ∀*i*]} efficiently controls the number of possible interventions and thus is more computationally efficient.

#### Proof

If action space A = {[*a*_*unaware*_, *a*_*ART*_]}:

We discuss this case in Remark 3.

If action space A = {[Δ*δ*_*i*_, 1 − Δ*ρ*_*i*_, ∀*i*]} the corresponding rates to simulate at every time-step would be *δ*_*t*_ − *δ*_*t+1*_ and *ρ*_*t*_ − *ρ*_*t+1*_:

Without loss of generality, we prove this Remark by showing that to generate one scenario equivalent of (*p*_*t − 1,U*_ − *p*_*t,U*_) = (*p*_*t,U*_ − *p*_*t+1,U*_) = 0, i.e., keeping proportion unaware constant over two consecutive time-steps, it would require evaluations of multiple combinations of *δ*_*t*_ and *δ*_*t+1*_ as the combination that generates (*p*_*t − 1,U*_ − *p*_*t,U*_) = (*p*_*t,U*_ − *p*_*t+1, U*_) = 0 would be dependent on system parameters at that time.

Writing equations for *δ*_*t*_ and *δ*_*t+1*_, setting (*p*_*t − 1,U*_ − *p*_*t,U*_) = (*p*_*t,U*_ − *p*_*t+1,U*_) = 0, and subtracting one from the other, we get

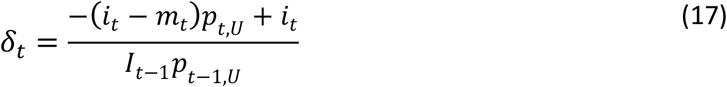

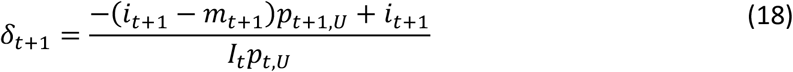

Subtracting equations (17) and (18) we get:

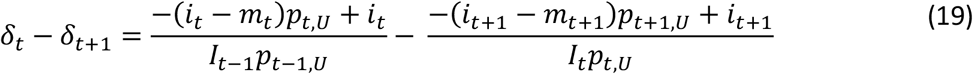

As (*p*_*t − 1,U*_ − *p*_*t,U*_) = 0 and (*p*_*t,U*_ − *p*_*t+1, U*_) = 0, we can set *p*_*t − 1,U*_ = *p*_*t,U*_ = *p*_*t+1,U*_ :

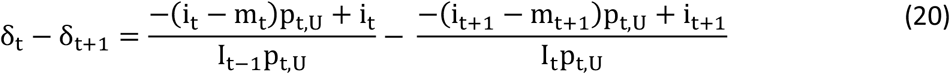

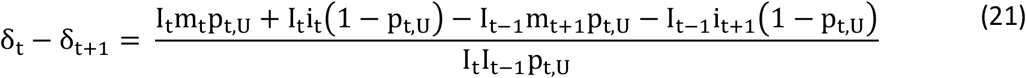

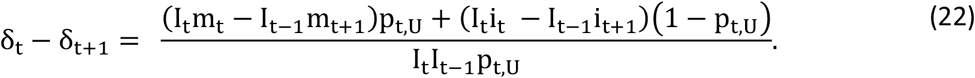

Therefore,(*p*_*t − 1,U*_ − *p*_*t,U*_) = (*p*_*t,U*_ − *p*_*t+1,U*_) = 0 could result from a range of diagnostic rate changes, *δ*_*t*_ − *δ*_*t+1*_ > 0, < 0, or = 0 depending on values of I_*t*_, m_*t*_, I_*t − 1*_, m_*t+1*_, *i*_*t*_, *a*nd *i*_*t+1*_. This implies that, if formulating an action as [Δ*δ*_*i*_, 1 − Δ*ρ*_*i*_, ∀*i*], a large subset of values for *δ*_*t*_ − *δ*_*t+1*_ should be evaluated as part of the action space. This is computationally expensive and, moreover, inefficient as many cases would result in an undesirable outcome.

Similarly, without loss of generality, to generate one scenario equivalent of (*p*_*t − 1,ART*_ − *p*_*t,ART*_) = (*p*_*t,ART*_ − *p*_*t+1,ART*_) = 0, i.e., keeping proportion on ART constant over two consecutive time-steps, it would require evaluations of multiple combinations of *ρ*_*t*_ *a*nd *ρ*_*t+1*_ as the combination that generates (*p*_*t − 1,ART*_ − *p*_*t,ART*_) = (*p*_*t,ART*_ − *p*_*t+1,ART*_) = 0 would be dependent on system parameters at that time.

Writing equations for *ρ*_*t*_ and *ρ*_*t+1*_, setting (*p*_*t − 1,ART*_ − *p*_*t,ART*_) = (*p*_*t,ART*_ − *p*_*t+1,ART*_) = 0, and subtracting one from the other, we get

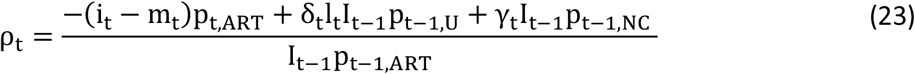

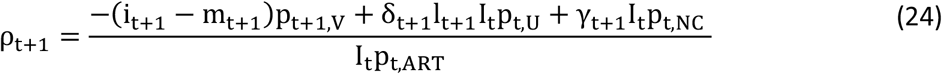

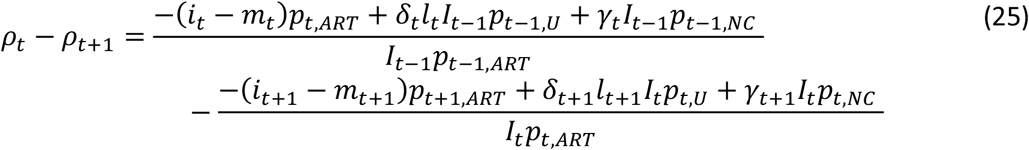

As (*p*_*t − 1,ART*_ − *p*_*t,ART*_) = 0 and (*p*_*t,ART*_ − *p*_*t+1,ART*_) = 0, we can set *p*_*t − 1,ART*_ = *p*_*t,ART*_ = *p*_*t+1,ART*_ :

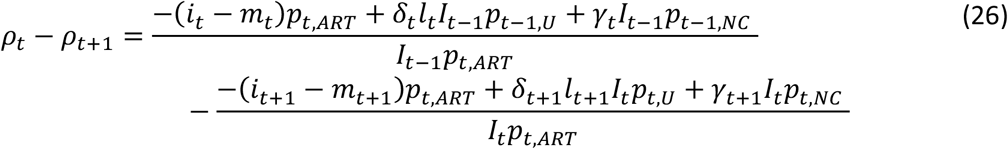

Rewriting with a common denominator,

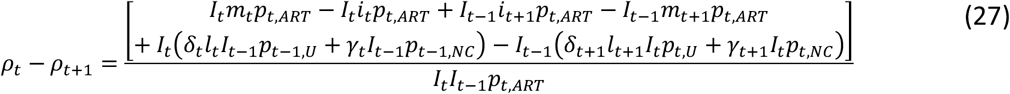

Rearranging the numerator,

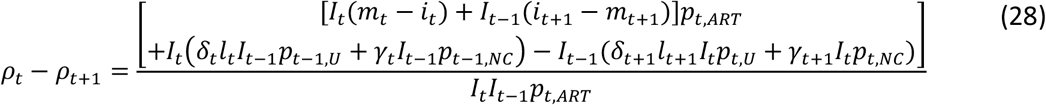

Therefore,(*p*_*t − 1,ART*_ − *p*_*t,ART*_) = (*p*_*t,ART*_ − *p*_*t+1,ART*_) = 0 could result from a range of drop-out rate changes, *ρ*_*t*_ − *ρ*_*t+1*_ > 0, < 0, or = 0, depending on values of *I*_*t*_, *m*_*t*_, *I*_*t − 1*_, *m*_*t+1*_, *i*_*t*_, *a*nd *i*_*t+1*_. This implies, if formulation an action as [Δ*δ*_*i*_, 1 − Δ*ρ*_*i*_, ∀*i*], a large subset of values for *ρ*_*t*_ − *ρ*_*t+1*_ should be evaluated as part of the action space. This is computationally expensive, and moreover inefficient, as most of the values would result in an undesirable outcome.

From Remarks 3 and 4, we can conclude that formulating the action space as A = {[δ_*i*_, 1 − ρ_*i*_, ∀*i* ∈ {HETs, MSM}]} would require evaluations of a large set of actions. For instance, if we use changes in testing and retention-in-care rates, we should consider both increase and decrease in these rates as both combinations can lead to higher proportion aware and proportion on ART. In this approach, choices would be combination of −25%, 0%, and 25% for testing rate and −20%, −10%, 0%, 10%, and 20% for retention-in-care (3 x 5 for HETs x 3 x 5 for MSM = 225). Or if we use changes in testing frequency and changes in retention-in-care rate as the action space, choices could be to test every 1, 2, 3, …, 10 years and retention-in-care rate can change by −20%, −10%, 0%, 10%, and 20% (10 x 5 for HETs x 10 x 5 for MSM = 2500). Problems with large action space generate issues of convergence, becoming infeasible to solve, and moreover, in this case, computationally inefficient, as a considerable portion of those values would result in (*p*_*t − 1,U*_ − *p*_*t,U*_) < 0, which is an undesirable outcome from a public health perspective (Remark 2). On the contrary, using action space A = {[*a*_*unaware,i*_, *a*_*ART,i*_∀*i*]} would naturally constrain the action space (to size 36 as discussed in paper section 2.1) by removing those actions that would result in undesirable outcomes, and is thus more efficient. Further, Remark 1 concludes that, for any given system state, solving for the optimal action [*a*_*unaware,i*_, *a*_*ART,i*_∀*i*] is equivalent to solving for optimal [δ_*i*_, 1 − ρ_*i*_, ∀*i* ∈ {HETs, MSM}].

### 4. Cost functions

We estimated the total cost of an action as the summation of the corresponding cost of the testing intervention program, retention-in-care intervention program, and treatment. The treatment costs are estimated in the PATH 2.0 simulation model by applying regimen-specific costs at the individual-level and are discussed elsewhere [2]. In section 2, for every action, we estimated the number of persons successfully intervened (numbers newly diagnosed and retained-in-care). Corresponding to those numbers, we discuss the estimation of the corresponding costs of testing and retention-in-care intervention programs in this section.

#### a. Estimation of HIV testing costs

In the estimation of testing costs, we make the following assumptions based on currently available data on testing behavior and testing intervention programs [4, 5, 6]. HIV testing programs can be conducted in clinical or non-clinical settings, each having its own fixed and variable costs [5, 6]. Fixed cost includes cost of clinics, other infrastructure, devices, equipment, staff, etc., while variable cost includes the cost per person tested. The marginal variable cost per additional person tested is a non-linear function of the proportion of the population tested and is influenced by the type of outreach program needed. Some people get tested voluntarily and incur only the cost of testing, while some get tested as a result of implementing an outreach intervention and thus incur additional costs of intervention [5]. Outreach intervention can include providers reaching out to the client’s community, delivering health information, reaching populations who have not been part of the traditional healthcare delivery system, HIV awareness campaigns, etc. [7]. The outreach intervention program is not 100% effective, meaning that not all outreached persons would get tested for HIV. To achieve the required number of persons tested under any action, outreach programs maybe be necessary. Under any given system state for proportion unaware, the corresponding cost of outreach is a non-linear function of the number of people outreached, i.e., the marginal cost to achieve one additional HIV-positive test increases as the proportion unaware decreases indicating more efforts would be needed to reach a larger portion of the population [8, 9]. The cost per person for persons testing positive is different from the cost per person for persons testing negative, as persons testing positive also undergo follow-up confirmation tests and additional care services for linkage to care [10, 11]. In accordance with current CDC recommendations, we assumed only persons with high risk are recommended for regular testing and applied testing costs for only these populations. We assumed that 6% of heterosexual females, 10% of heterosexual males, and all MSM are high-risk [12, 13, 14].

We estimate the cost of testing corresponding to an action *a* as follows:

Let

*r*_*t,a*_ be the number of persons testing positive at time step t under action *a*,

*n*_*t,a*_ be the number of persons testing negative at time step t under action *a*,

*x*_*t,a*_ be the number of persons reached through an outreach testing intervention program at time step t under action *a*,

*Y*_v_ be the variable cost per person testing positive,

*X*_v_ be the variable cost per person testing negative,

*O*_v_ be the variable cost per person outreached through an outreach testing intervention program,

*X*_f − cl,*a*_ be the total fixed cost of testing in a clinical setting under action *a*,

*X*_f − ncl,*a*_ be the total fixed cost of testing in a non-clinical setting under action *a*,

*X*_f − o,*a*_ be the total fixed cost of implementing an outreach intervention program under action *a*, and

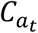 be the total cost of testing under action *a*.

Then, we can calculate the total cost of testing action as

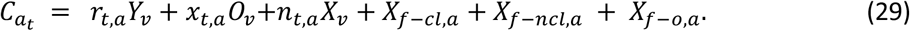

Note that, *r*_*t,a*_ is the number of persons successfully intervened, i.e., the persons newly diagnosed as estimated in Appendix section 2. To achieve this, a larger number of persons must have tested negative (*n*_*t,a*_), and further, a larger number of persons must have been reached through an intervention (*x*_*t,a*_) as it is not necessary that all who are intervened would take a HIV test. Therefore, using our estimates of *r*_*t,a*_ and based on other data from the literature on the effectiveness of interventions programs, we estimate *n*_*t,a*_ and *x*_*t,a*_. Furthermore, as the unit costs for persons in each category (*r*_*t,a*_, *n*_*t,a*_, or *x*_*t,a*_) are also likely different, we estimate the unit costs for each category (*Y*_v_, *O*_v_, *X*_v_, respectively) in addition to the fixed costs using data from the literature. Below, we discuss the estimation of each of these components on the right-hand side of the (29).

**Estimation of *r***_***t***,***a***_**-**Number of persons testing positive at time step t under action *a*: Note that, this is the number of persons successfully intervened, i.e., the persons newly diagnosed as estimated in Appendix section 2, i.e.,

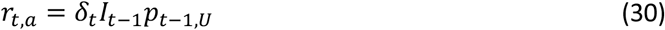

where

*δ*_*t*_ is the rate of HIV diagnosis under action *a*,

*I*_*t*_ is the number of infected persons at time *t*, and

*p*_*t*,U_ is the proportion of infected persons in stage unaware at time *t*.

Estimation of *δ*_*t*_ was discussed earlier, and *I*_*t* − 1_ and *p*_*t* − 1,U_ are simulated in the PATH 2.0 model.

**Estimation of *x***_***t***,***a***_ -Number of people outreached at time step t under action *a*:

Let

*μ* be the rate of diagnosis through voluntary testing,

*θ* be the effectiveness of an outreach intervention program,

*φ* be the proportion of people testing positive for HIV among those tested through an outreach intervention program, and

*N*_*t*_ be the total population at time *t*.

Then we can write,

*μI*_*t* − 1_*p*_*t* − 1,U_ as the number of persons diagnosed through voluntary testing, and *x*_*t,a*_*θφ* as the number of persons diagnosed through an outreach testing intervention program,

i.e.,

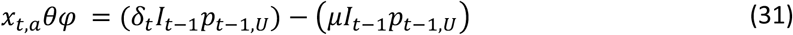

And thus, estimate the total number of people outreached as

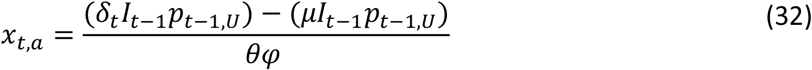

We calculate the unknown terms on the right-hand side of the above equation as follows, 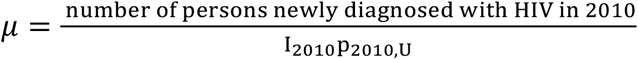 by assuming that the rate of HIV diagnosis in the year 2010, which is just prior to the implementation of the first HIV national strategic plan [15], is the rate of diagnosis through voluntary testing and that it remains the same for future years,

*θ* = 30% an assumption taken from [6], and

*φ* = 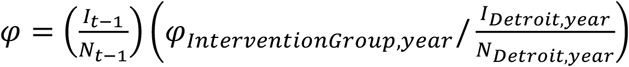,

where *φ*_*InterventionGroup,year*_ = 0.0153 is the proportion of persons testing positive in the outreach intervention program, (Shrestha Clark) and 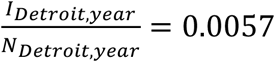 is the prevalence of HIV in Detroit at the time the study was conducted^1^.

**Estimation of *n***_***t***,***a***_**-**Number of persons testing negative at time step t under action *a*: Following from the previous subsection, we can write

*x*_*t,a*_*θ*(1 − *φ*) as the number of persons testing negative for HIV among those tested through an outreach intervention program, and

*μ*(*N*_*t*_ − *I*_*t*− 1_*p*_*t*− 1,U_ − *x*_*t,a*_) as the number of persons testing negative for HIV among those who voluntarily get tested.

Thus, we estimate the total number of HIV negative cases as:

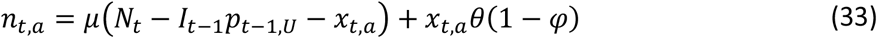

The estimation of all parameters on the right side of the equation has been discussed earlier.

**Estimation of *X***_***v***_ **-**Variable cost per person testing negative:

Let

*c*_r,n_ be the rapid test cost for an HIV-negative case,

*c*_c,n_ be the conventional test cost for an HIV-negative case,

*n*_n_ be the notification cost for an HIV-negative case,

*c*_*a*dd_ be the additional cost per person performed in a non-clinical setting,

*τ* be the proportion rapid test, and

*α* be the proportion tested in clinical setting.

We can then estimate the variable cost per person for a HIV negative case as

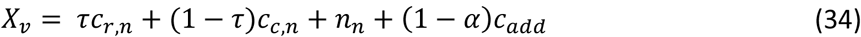

For the parameters on the right-hand side of the equation, we use estimates from the literature [6] and are summarized in Table A1.

**Estimation of *Y***_***v***_ – Variable cost per person testing positive:

Let

*c*_r,p_ be the rapid test cost for an HIV-positive case,

*c*_c,p_ be the conventional test cost for an HIV-positive case,

*n*_r,p_ be the rapid test notification cost for an HIV-positive case,

*n*_c,p_ be the conventional test notification cost for an HIV-positive case, and

*c*_cnf_ be the confirmatory cost.

Hence, we estimate the variable cost per person for a HIV positive case as follows:

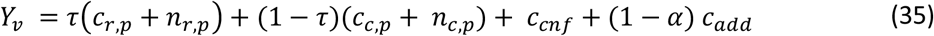

For the parameters on the right-hand side of the equation, we use estimates from the literature [6] and are summarized in Table A1.

**Estimation of *O***_***v***_-Variable cost per person outreached:

As the proportion of HIV-infected persons unaware of their infection decreases, the marginal cost of reaching one additional HIV-positive person increases. Therefore, we formulate the variable cost of outreach intervention as a non-linear function of increment in the proportion of people outreached relative to the number of people outreached in the base year with respect to the total population.

Let

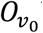be the base year variable cost for outreaching *x*_0_ people among total population in the base year (2015),

*x*_0_ be the number of people outreached in the base year estimated using data in (Shrestha Clark) with outreach intervention effectiveness of 30% [6],

*Δx* be the increment in the number of populations outreached from the base year with respect to total population at time t, calculated as 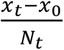, and

*w* be the coefficient of a non-linear variable cost function.

Then per person variable cost of *Δx* increment in outreaching *i*s:

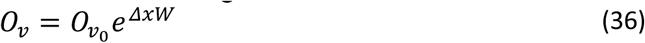

Derivation:

We assume that *Δx* increase in the number of people outreached imposes extra cost of *O*_v_*wΔx* and estimate per person outreach cost as,

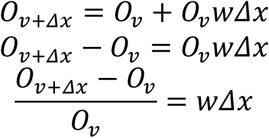

Integrating on both sides,

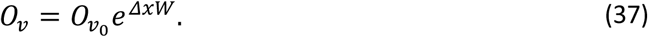

For the parameters on the right-hand side of the equation, we use estimates from the literature and are summarized in

Table A2.

**Estimation of *X***_***f***− ***cl***,***a***_ – Total fixed cost of testing in a clinical setting corresponding to action *a*:

We assume that infected and uninfected persons share the same fixed costs in a clinical setting:

Let

*α* be the proportion of people testing in a clinical setting,

*m*_*cl*_ be capacity of a clinic (we use an average estimate for clinic capacity),

*f*_*c*_ be the total fixed cost per clinic with m_cl_ capacity, and

*X*_*f− cl,a*_ be the total clinical fixed cost.

Then the total fixed cost for testing α(r_*t,a*_ + n_*t,a*_) number of people in a clinical setting is estimated as follows:

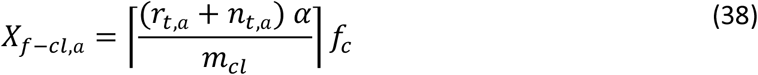

For the parameters on the right-hand side of the equation, which were not discussed earlier, we use estimates from the literature [11, 10, 5], and are summarized in Table A1.

**Estimation of *X***_***f***− ***ncl***,***a***_ – Total fixed cost of testing in a non-clinical setting corresponding to action *a*:

We assume that infected and uninfected persons share the same fixed costs in non-clinical testing.

Let

m_nc_ be the average capacity of a non-clinical setting and

f_nc_ be the total fixed cost per non-clinic setting with capacity m_nc_.

Then total fixed cost to test (1 − *α*)(*r*_*t,a*_ + *n*_*t,a*_) people in a non-clinical setting is estimated as follow:

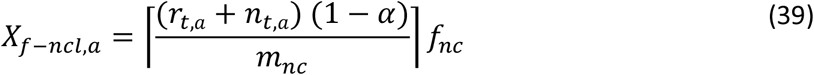

For the parameters on the right-hand side of the equation that were not discussed earlier, we use estimates from the literature [10, 11, 16, 17], and are summarized in Table A1.

**Estimation of *X***_***f***− ***o***,***a***_-Fixed cost of outreach intervention:

Let

*m*_o_ be the capacity of outreach intervention, and

*f*_o_ be the total fixed cost of outreaching *x*_*t*_ people.

Then the total fixed cost to outreach *x*_*t,a*_ number of people is estimated as

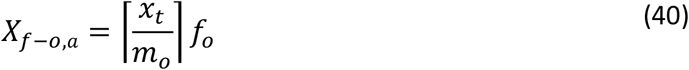

For the parameters on the right-hand side of the equation that were not discussed earlier, we use estimates from the literature [5] and are summarized in Table A2.

**Table A1:** Cost Components of HIV Positive and Negative Testing^2^

| Input | Value | Source |
| --- | --- | --- |
| Testing cost, including screen and confirmatory test |  |  |
| HIV-uninfected individual, rapid screen | \$22.13 (2012\$) | Based on cost components from [10, 11, 16]. |
| HIV-uninfected individual, conventional screen | Before 2016: \$8.24 (2012\$)<br>2016+: \$10.36 (2012\$) | Based on cost components from [11, 16, 17]. |
| HIV-infected individual, rapid screen | Before 2016: \$86.70 (2012\$)<br>2016+: \$78.80 (2012\$) | Based on cost components from [10, 11, 16, 17] and adjusted to 2012\$. Assumes a repeat screen and a Western blot confirmatory test before 2016; a Geenius HIV 1/2 confirmatory screen is used in 2016 and after. |
| HIV-infected individual, conventional screen | Before 2016: \$60.02 (2012\$)<br>2016+: \$58.91 (2012\$) | Based on cost components from [11, 16, 17] and adjusted to 2012\$. Assumed Western blot confirmatory test before 2016, and a Geenius HIV 1/2 confirmatory screen in 2016 and after. |
| NAT<sup>3</sup>, applied for discrepant Western blot confirmatory test | \$160.07 (2012\$) | [17] |
| Additional cost per test performed in non-clinical (vs. clinical) setting | \$52.66 (2005\$) | [5] |
| Notification costs |  |  |
| HIV-uninfected | \$0.45 (2009\$) | [16] |
| HIV-infected, conventional screen | \$5.88 (2009\$) | [16] |
| HIV-infected, rapid screen | \$10.86 (2009\$) | [16] |
| Outreach cost per test (when applied) | \$13.67 (2005\$) | [5] |

<sup>2</sup> The component of testing costs are taken from HOPE model technical report [6].
<sup>3</sup> Note: NAT = HIV nucleic acid amplification test

**Table A2.** Summary of Parameters and Values Used in Testing and Outreach Intervention

| Notation | Description | Value(s) | Reference |
| --- | --- | --- | --- |
| $O_{v_0}$ | Base year variable cost for outreaching $x_0$ people | \$16.59 (2015\$) | [5] |
| $\Delta x$ | Increment in the number of populations outreached from the base year | | Calculated in PATH |
| $W$ | coefficient of variable cost | 0.1, 0.2, 0.3 | Assumption |
| $m_{cl}, m_{nc}$ | Clinical/ non-clinical capacity | 1000 | Assumption |
| $f_c$ | Total fixed cost per clinic with $m_{cl}$ capacity | \$56379 (2015\$) | [5] |
| $f_{nc}$ | Total fixed cost per clinic with $m_{cl}$ capacity | \$64851 (2015\$) | [5] |

#### b. Estimation of retention-in-care cost

We assume that the required proportion on ART under any action (*a*) can be achieved by implementing retention-in-care programs that ensure patients remain in care and consistently take antiretroviral therapy treatment to achieve viral load suppression [18]. Retention-in-care programs could include a combination of a face-to-face meeting with patients at primary care visits, brief interim phone contacts between appointments, appointment reminders, and missed-visit calls [18]. We assume retention-in-care programs could include fixed and variable costs. Fixed costs include the cost of office space, durable items such as computers, printers, telephones, etc. And variable costs include staff time spent on management of patients and personal contact with patients [18]. We assume the variable cost per person is a non-linear function of the proportion of people retained in care, i.e., the marginal cost to achieve one additional person retained in care increases as the proportion on ART increases indicating more efforts would be needed to retain a larger number of people in care.

We estimate the cost of retention-in-care corresponding to an action as follows.

Let

*d*_*t,a*_ be the number of persons retained in care at time step t under action *a*;

*R*_v_ be the variable cost per person retained in care,

*E*_f,*a*_ be the total fixed cost of retaining in care under action *a*, and

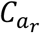 be the total cost of retention in care under action *a*.

Then, we can calculate the total cost retention in care action as:

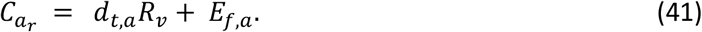

Below we discuss the estimation of each component on the right-hand side of the equation.

**Estimation of *d***_***t***,*a*_**-**Number of people retained in care:

Note that, this is the number of persons successfully intervened (number retained-in-care) and estimated in Appendix section 2, i.e.,

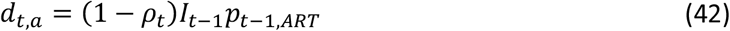

The parameters on the right-hand side of the equation are calculated in the PATH 2.0 simulation model.

**Estimation of *R***_***v***_**-**Retention-in-care variable costs:

Let

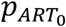 be the proportion on ART in the base year 2015, which is calculated in PATH,

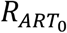 be the base year variable cost of achieving 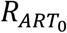,

*Δp*_*ART*_ is the increment in the proportion on ART with respect to the base year, and

*Y* is the coefficient of the non-linear variable cost function.

Similar to testing outreach variable cost, we calculate the retention-in-care variable cost as follows:

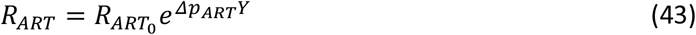

For the parameters on the right hand-side of the equation that were not discussed earlier, we use estimates from the literature [18] and are summarized in Table A3.

**Estimation of *E***_***f***,***a***_**-**Retention-in-care fixed costs:

Let

*m*_r_ be the retention-in-care intervention setting capacity, and

*f*_*r*_ be the fixed cost per retention-in-care intervention program (with capacity *m*_*r*_).

We estimate the total fixed cost for retaining *d*_*t,a*_ number of people in care as:

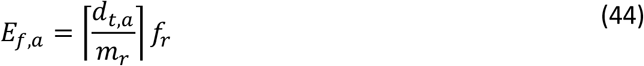

For the parameters on the right hand-side of the equation that were not discussed earlier, we use estimates from the literature (Shrestha Gardner) and are summarized in Table A3.

**Table A3:** Summary of Parameters Used in Retention-in-care Intervention

| Notation | Description | Value(s) | Reference |
| --- | --- | --- | --- |
| $p_{ART_0}$ | Proportion on ART at time step 0 | | Calculated in PATH |
| $R_{ART_0}$ | Base year variable cost of achieving $p_{ART_0}$ | \$117 (2015\$)<br>\$235 (2015\$)<br>\$300 (2015\$) | Low, medium, and high retention-in-care costs [18] |
| $\Delta p_{ART}$ | Increment in the proportion on ART | | Calculated in PATH |
| $Y$ | Coefficient of variable cost | 0.1, 0.2, 0.3 | Assumption |
| $m_r$ | Retention-in-care intervention program capacity | 500 | Assumption based on [18] |
| $f_r$ | Fixed cost per retention-in-care intervention program with capacity $m_r$ | \$17977 (2015\$)<br>\$22708 (2015\$)<br>\$29330 (2015\$) | Low, medium, and high retention-in-care costs [18] |

### 5. Q-Learning algorithm

**Figure A3:**
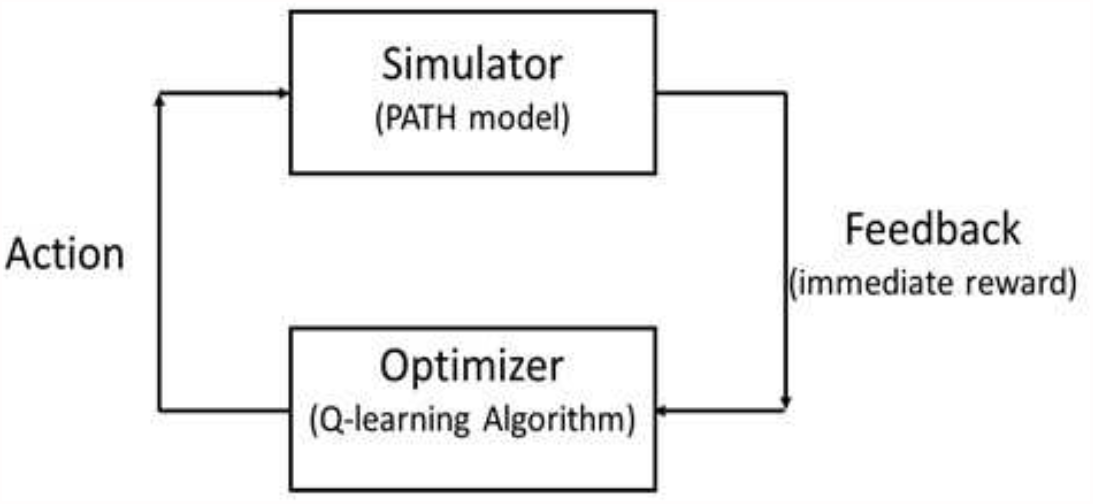
Schematic of the reinforcement learning (RL) algorithm. The RL algorithm takes action, and it is fed into the simulator. The simulator simulates the action and returns an immediate reward to the algorithm to update the following action accordingly.

**Table A4:**
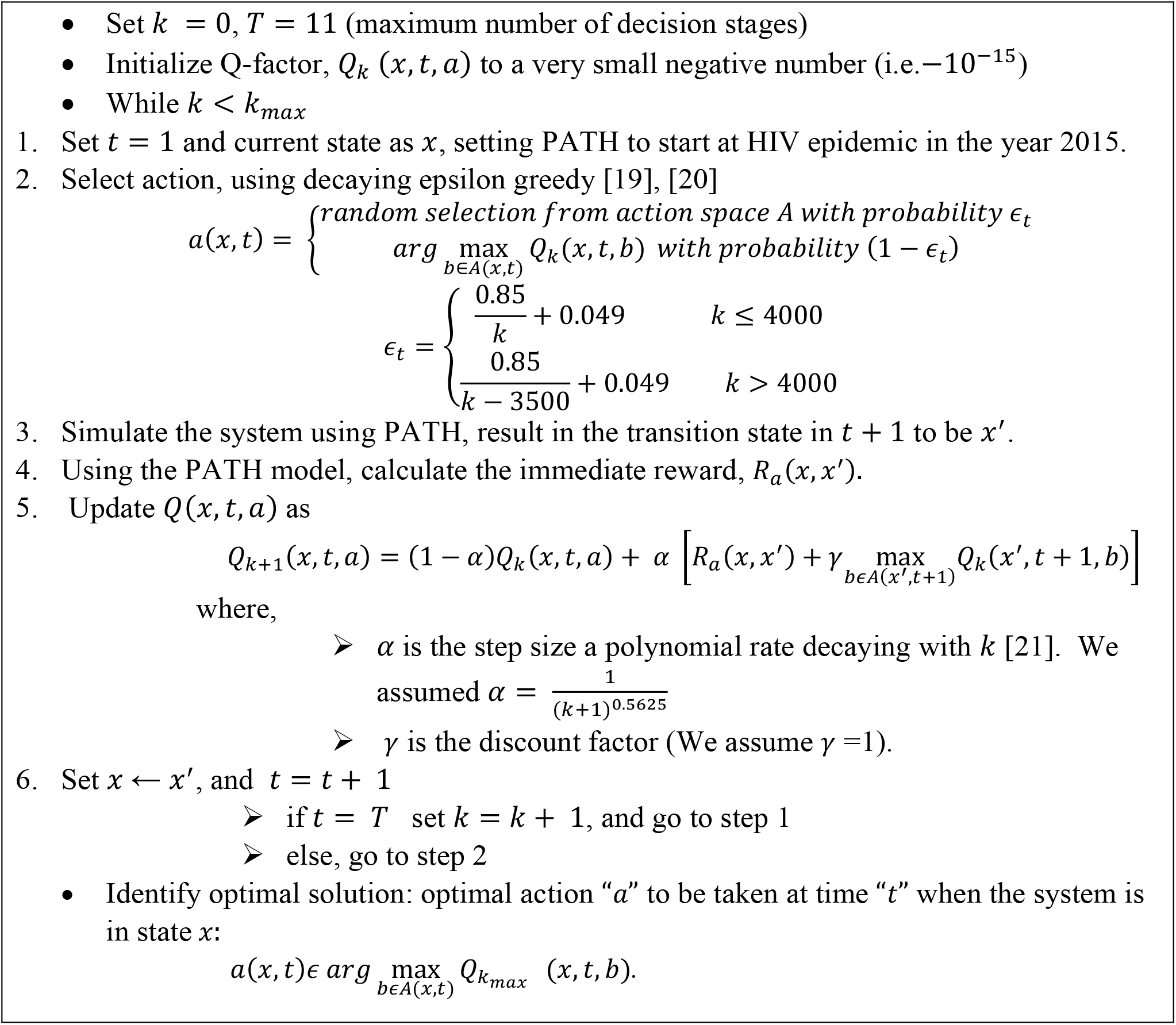
Finite-horizon Q-learning Algorithm to Identify Optimal Solutions to the Markov Decision Process [1].

### 7. Testing convergence of the Q-learning reinforcement algorithm

In this section, we present the uncertainty in results, specifically, the uncertainty in the optimal policy [*a*_*unaware,HET*_, *a*_*ART,HET*_, *a*_*unaware,MSM*_, *a*_*ART,MSM*_] and the uncertainty in values of testing and retention-in-care rates [*δ*, 1 − *ρ*], corresponding to each scenario for different values of Q-learning training iterations *k*_m*a*x_ (2000, 3000, 4000, and 5000). An algorithm is said to have converged if, through the iterative search process, it has reached a local optima, i.e., successfully solved for an optimal combination of testing and retention-in-care rates. If the number of iterations is not sufficiently large, there is a risk that the algorithm is terminated before convergence. The ideal number is typically determined through experimentation.

Further, there could be multiple local optima, i.e., multiple policies could yield similar total rewards, and because of the stochastic nature of the epidemic system, the optimal policy could be a range rather than a point estimate. Therefore, we ran the model for varying number of iterations, 2000, 3000, 4000, and 5000, and compared the corresponding total rewards, to ensure convergence and obtain the uncertainty range in optimal policies (Figures Figure A4,Figure A7Figure A10 for each of the three cost functions). Note also that the training with *k*_*max*_ = 5000 has an additional exploration after 4000 iterations (defined by the epsilon-greedy action selection defined in main manuscript). The relative difference in the total population costs between the varying iterations were at most 2% in each cost function evaluated, suggesting convergence (Figures Figure A6,Figure A9, and Figure A12 for each of the three cost functions). The changes in new infections and the number of people with HIV (PWH) were also minimal (Figures Figure A5,Figure A8, andFigure A11 for each of the three cost functions). The corresponding optimal policies differed slightly, more so in future years than earlier years, suggesting stochastic uncertainty as the model projects further into the future (Figures Figure A4Figure A7Figure A10 for each of the three cost functions). Therefore, in Results of main paper, we present the range of optimal policies across these iterations as the uncertainty range.

**Figure A4:**
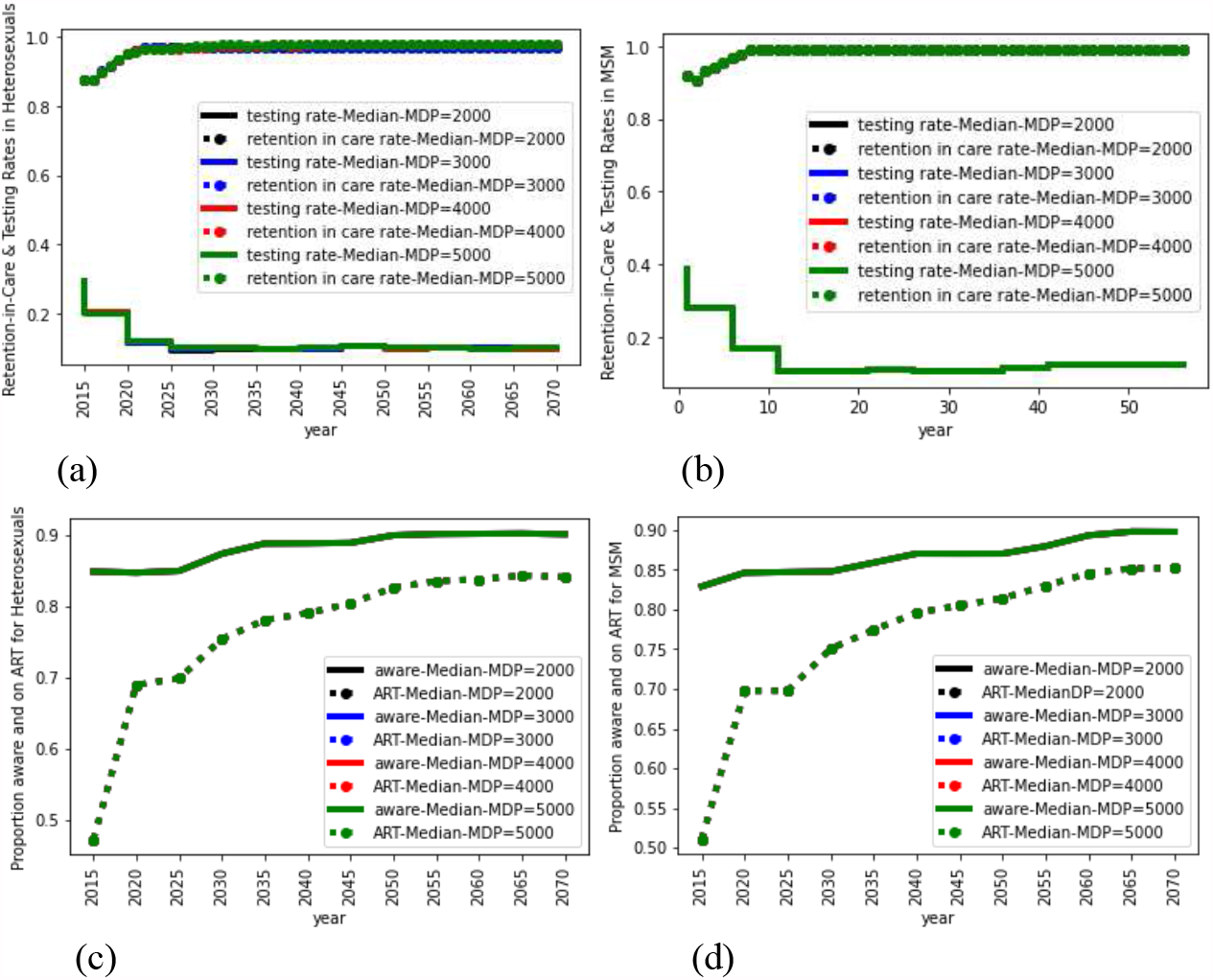
Medium Cost Function: Combination of optimal policy (testing and retention-in-care rates) for heterosexuals (a) and MSM (b), and corresponding proportion of aware and on ART for heterosexuals (c) and MSM (d) for MDP iterations of 2000 (black), 3000 (blue), 4000 (red), and 5000 (green). Results are an average of 100 runs.

**Figure A5:**
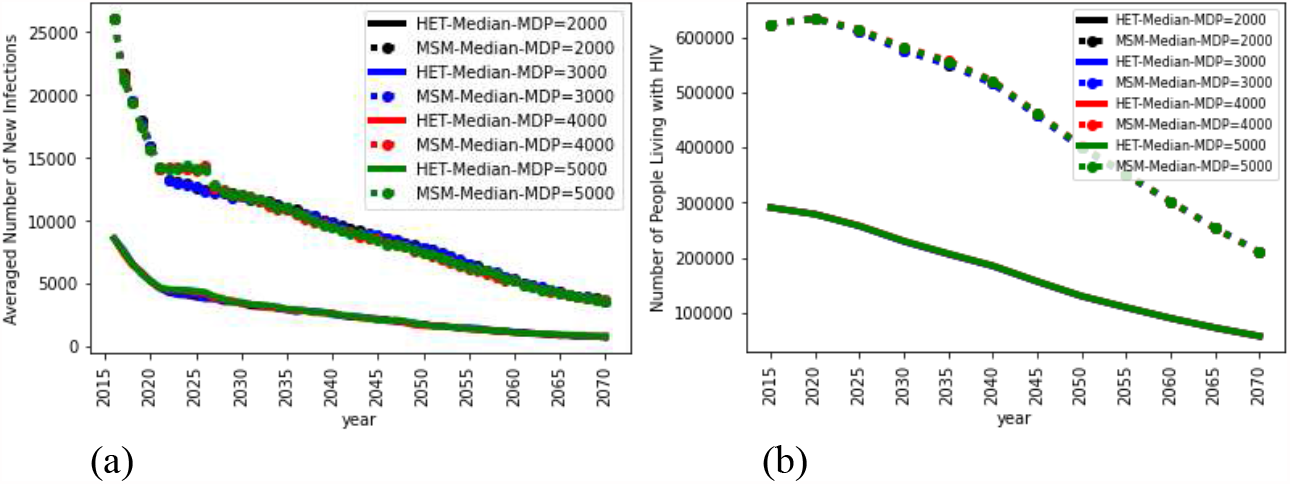
Median Cost Function: Impact of implementing a combination of optimal policy on the number of new infections (a) and the number of people living with HIV (b) for heterosexuals (solid lines) and MSM (dashed lines) for MDP iterations of 2000 (black), 3000 (blue), 4000 (red), and 5000 (green). Results are an average of 100 runs.

**Figure A6:**
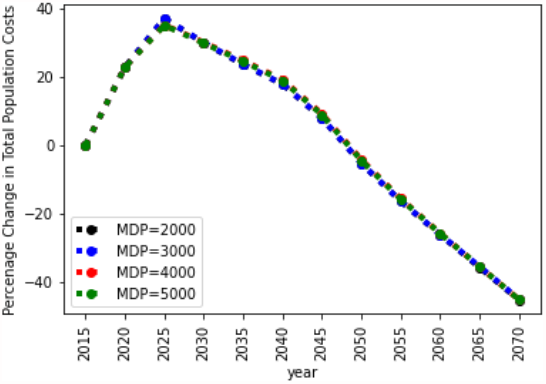
Median Cost Function: Percentage increment in total population cost of implementing optimal policy for MDP iterations of 2000 (black), 3000 (blue), 4000 (red), and 5000 (green). Results are an average of 100 runs.

**Figure A7:**
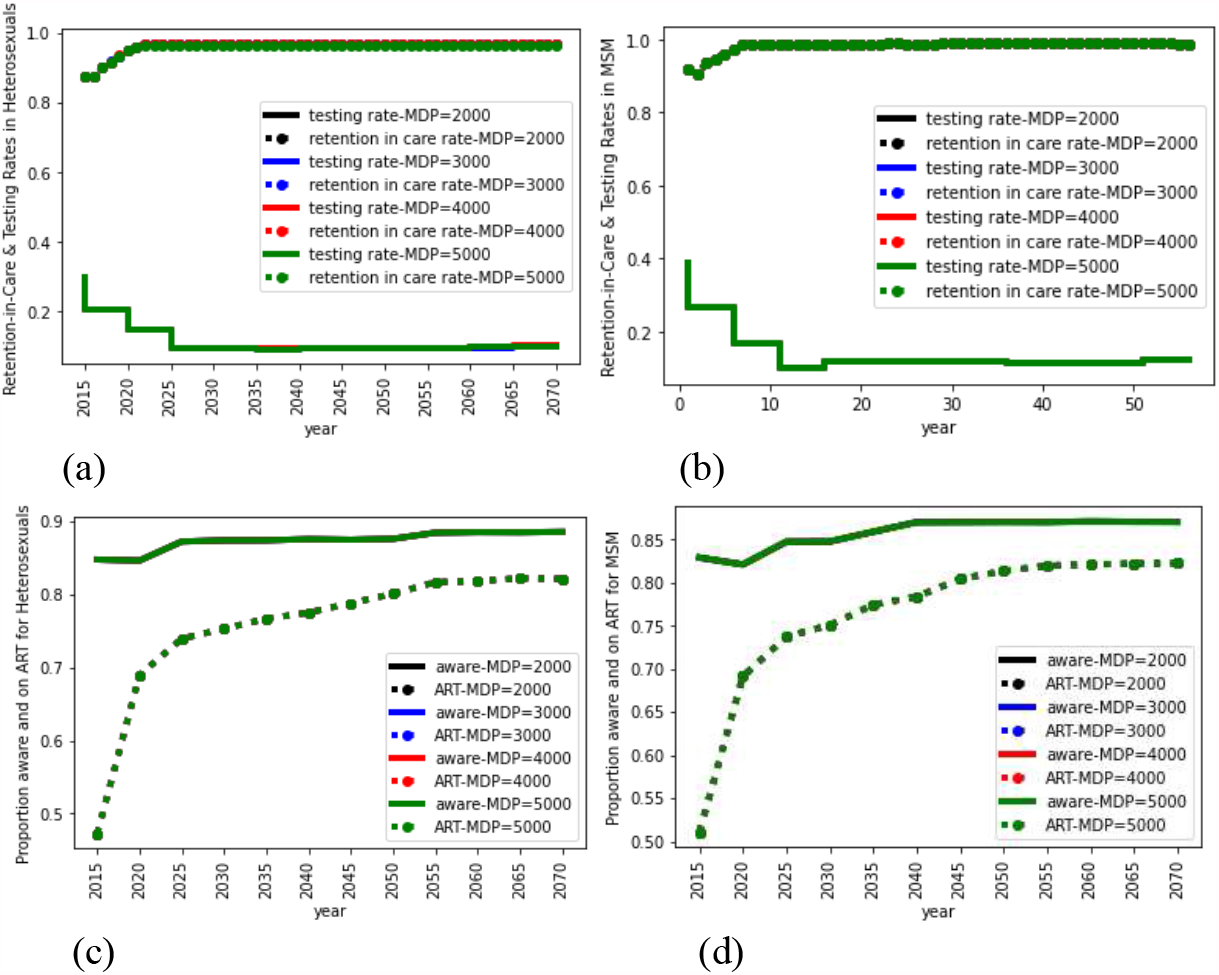
HTLR Cost Function: Combination of optimal policy (testing and retention-in-care rates) for heterosexuals (a) and MSM (b), and corresponding proportion of aware and on ART for heterosexuals (c) and MSM (d) for MDP iterations of 2000 (black), 3000 (blue), 4000 (red), and 5000 (green). Results are an average of 100 runs.

**Figure A8:**
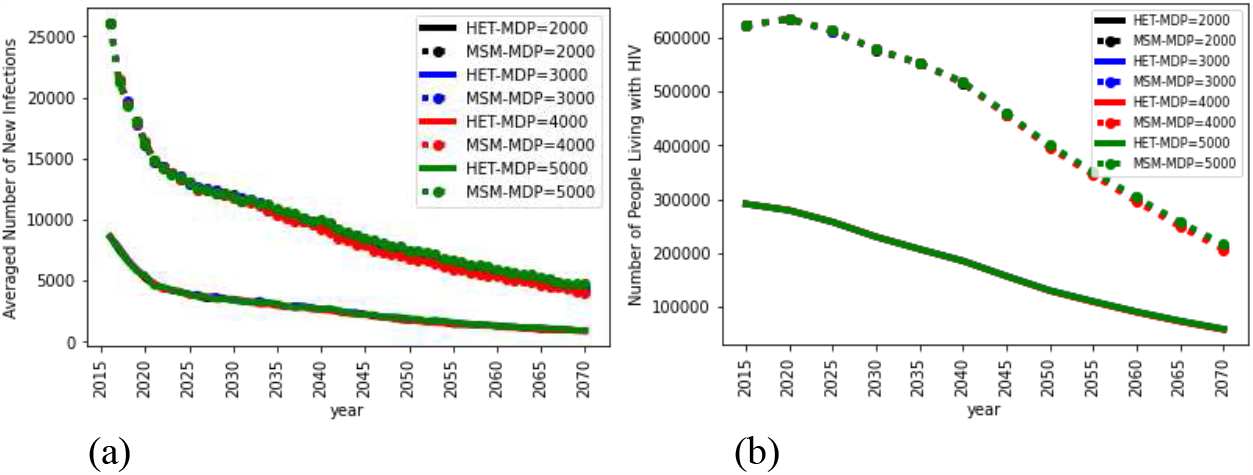
HTLR Cost Function: Impact of implementing combination of optimal policy on the number of new infections (a) and the number of people living with HIV (b) for heterosexuals (solid lines) and MSM (dashed lines) for MDP iterations of 2000 (black), 3000 (blue), 4000 (red), and 5000 (green).

**Figure A9:**
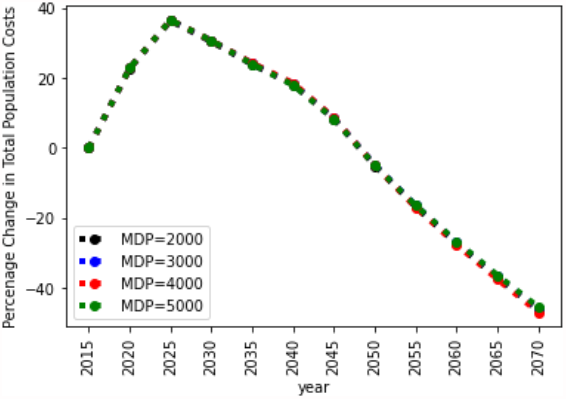
HTLR Cost Function: Percentage increment in total population cost of implementing optimal policy for MDP iterations of 2000 (black), 3000 (blue), 4000 (red), and 5000 (green). Results are an average of 100 runs. Results are an average of 100 runs.

**Figure A10:**
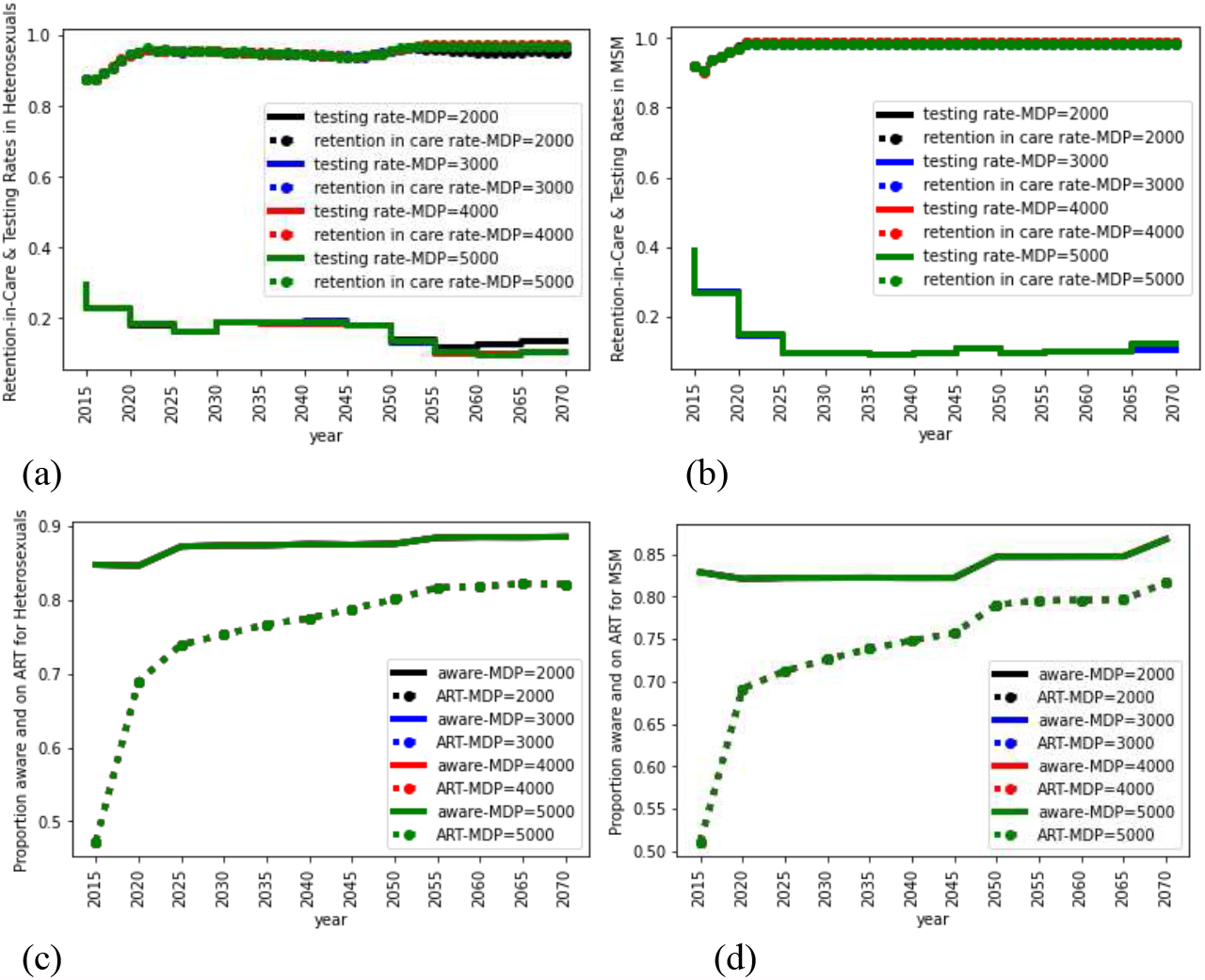
LTHR Cost Function: Combination of optimal policy (testing and retention-in-care) for heterosexuals (a) and MSM (b), and corresponding proportion of aware and on ART for heterosexuals (c) and MSM (d) for MDP iterations of 2000 (black), 3000 (blue), 4000 (red), and 5000 (green). Results are an average of 100 runs.

**Figure A11:**
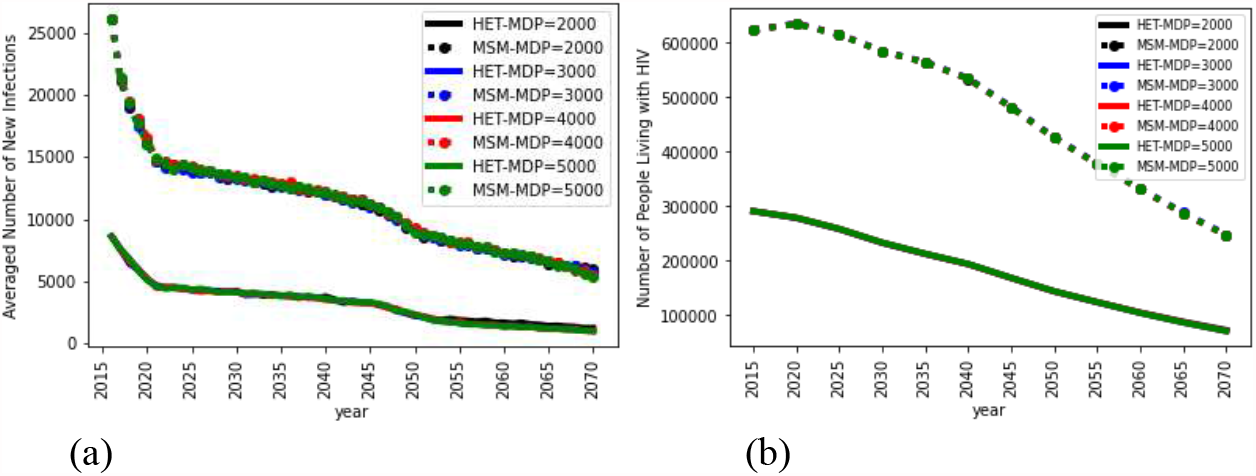
LTHR Cost Function: Impact of implementing combination of optimal policy on the number of new infections (a) and the number of people living with HIV (b) for heterosexuals (solid lines) and MSM (dashed lines) for MDP iterations of 2000 (black), 3000 (blue), 4000 (red), and 5000 (green). Results are an average of 100 runs.

**Figure A12:**
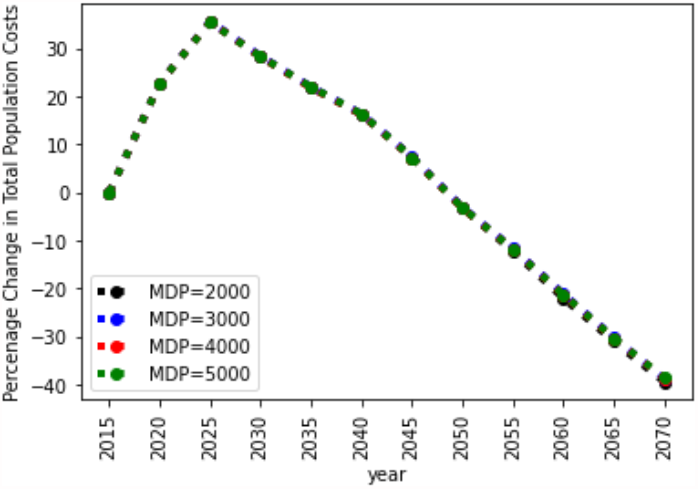
LTHR Cost Function: Percentage increment in total population cost of implementing optimal policy for MDP iterations of 2000 (black), 3000 (blue), 4000 (red), and 5000 (green). Results are an average of 100 runs.

### 8. Evaluating the optimality of the counter-intuitive results under the LTHR MSM scenario

As discussed in the manuscript, comparing across the cost function assumptions, the optimal rates were generally intuitive for heterosexuals, highest testing and lowest retention-in-care in LTHR (which assumed lowest unit cost for testing and highest unit cost for retention-in-care) and lowest testing and highest retention-in-care rates in HTLR, though the differences in retention-in-care rates were modest (paper Figure 1a). For MSM, however, though the model suggested optimal rates were similar in all three cost functions, it counter-intuitively suggested a slightly lower testing rate in LTHR compared to Median and HTLR. It suggested to instead spend those resources on maintaining retention-in-care rates at the level of Median and HTLR (paper Figure 1b), such that the proportion of MSM on ART in LTHR, though lower than in Median and HTLR, was higher than the proportion of heterosexuals on ART in LTHR (paper Figure 1d). The optimality of this counter-intuitive strategy in MSM was evaluated by a counterfactual simulation run using the optimal LTHR strategy of heterosexuals for both heterosexuals and MSM (FigureFigure A13). The number of new infections, PWH, and costs were higher in the counterfactual simulation, confirming the optimality of the policy (Figures Figure A14 and Figure A15). The results of this counterfactual run also suggest that the reasoning behind the counter-intuitive strategy in MSM is likely because of the higher prevalence of HIV in MSM, i.e., lowering the spending on retention-in-care would lead to more infections than lowering the spending on testing. Note also in this counterfactual run, the results for heterosexuals also change over time compared to the original (though they both start with the same optimal strategy) because of the dynamics of contact mixing between MSM and heterosexuals over time and the functionality of the model to correct for those dynamic changes and find a new optimal. A similar counterfactual run of using the MSM optimal strategy on the heterosexual risk group generated similar findings.

**Figure A13:**
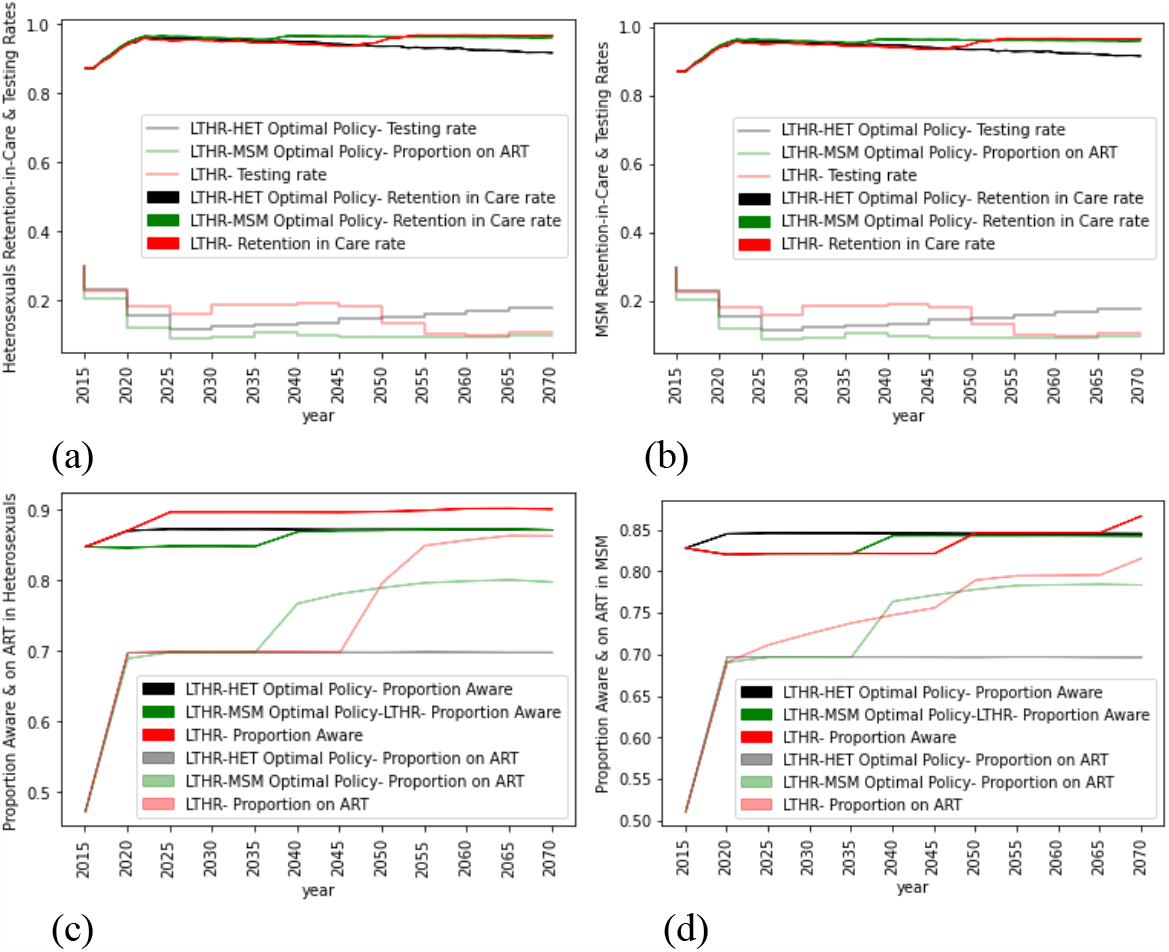
Top: Optimal combination of testing rate (a) and retention-in-care rate (b). Bottom: Corresponding proportion aware (c) and proportion on ART (d).

**Figure A14:**
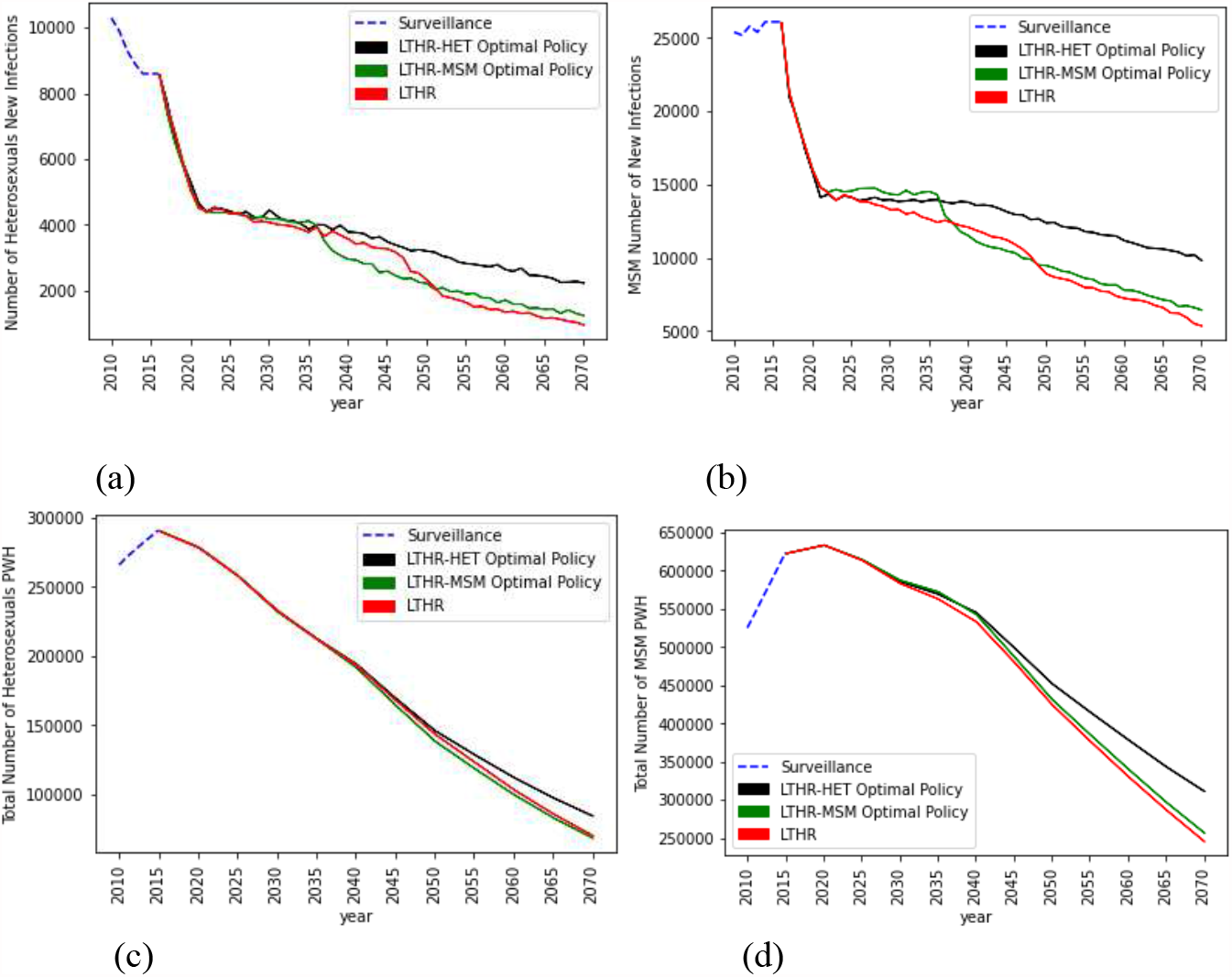
Top: Number of new infections for heterosexuals (a) and MSM (b). Bottom: Number of people living with HIV for heterosexuals (a) and MSM (b).

**Figure A15:**
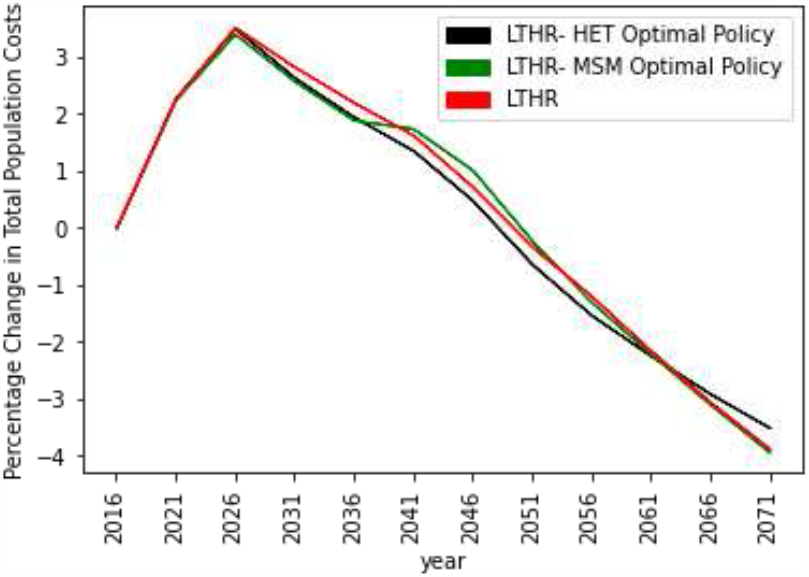
Comparing percentage change in total population cost of LTHR cost function (red) with heterosexual LTHR cost function applied to both risk groups (black) and with MSM LTHR cost function applied to both risk groups (green) all with MDP=5000.

## Footnotes

1 This equation assumes that the outreach intervention program would be targeted towards higher-risk individuals and thus the proportion testing positive would be higher than the overall prevalence in the population, as evident by the data (presented above) in this study [5].

2 The component of testing costs are taken from HOPE model technical report [6].

3 Note: NAT = HIV nucleic acid amplification test

